# Identifying SARS-CoV-2 Lineages that Share the Same Relative Effective Reproduction Numbers

**DOI:** 10.64898/2026.04.22.26351531

**Authors:** Richard Musonda, Koichi Ito, Ryosuke Omori, Kimihito Ito

## Abstract

The severe acute respiratory syndrome coronavirus 2 (SARS-CoV-2) has continuously evolved since its emergence in the human population in 2019. As of 1st August 2025, more than 1,700 Omicron subvariants have been designated by the Pango nomenclature system. The Pango nomenclature system designates a new lineage based on genetic and epidemiological information of SARS-CoV-2 strains. However, there is a possibility that strains that have similar genetic backgrounds and the same phenotype are given different Pango lineage names. In this paper, we propose a new algorithm, called FindPart-*w*, which can identify groups of viral lineages that share the same relative effective reproduction numbers. We introduced a new lineage replacement model, called the constrained RelRe model, which constrains groups of lineages to have the same relative effective reproduction numbers. The FindPart-*w* algorithm searches the equality constraints that minimise the Akaike Information Criterion of constrained RelRe models. Using hypothetical observation count data created by simulation, we found that the FindPart-*w* algorithm can identify groups of lineages having the same relative effective reproduction number in a practical computational time. Applying FindPart-*w* to actual real-world data of time-stamped lineage counts from the United States, we found that the Pango lineage nomenclature system may have given different lineage names to SARS-CoV-2 strains even if they have the same relative effective reproduction number and similar genetic backgrounds. In conclusion, this study showed that viruses that had the same relative effective reproduction number were identifiable from temporal count data of viral sequences. These findings will contribute to the future development of lineage designation systems that consider both genetic backgrounds and transmissibilities of lineages.

## Introduction

Since the emergence of the severe acute respiratory syndrome coronavirus 2 (SARS-CoV-2) in the human population in 2019 (1), this virus has continuously evolved by accumulating mutations. A group of viral strains that share similar genetic background and the same phenotypic properties is called a variant. When a variant is more transmissible in the human population than other variants, the frequency of the variant increases over time through natural selection (2). So far, five SARS-CoV-2 variants, Alpha, Beta, Gamma, Delta, and Omicron, have been designated as variants of concern (VOC) by the World Health Organization (WHO) (3). Of these, the most transmissible variant is Omicron (4). WHO designated Omicron as a VOC on November 26, 2021 (5), and Omicron became dominant globally within four weeks (6). Since then, subvariants of Omicron having immune evasion properties have been continuously emerging until now (7).

A variant can be decomposed into finer units called lineages and clades, and a name is assigned to each lineage or clade by a nomenclature system. Giving names to lineages and clades allows us to identify a group of viruses, and having a good nomenclature system is essential for effective communication, especially among the research community (3).

Several nomenclature systems have been developed and used so far. Each nomenclature system applies its own criteria to designate a new lineage or clade. The most widely used nomenclature system is the Pango nomenclature system (8). The Pango system designates a new lineage based on shared nucleotide differences from an ancestral lineage, phylogenetic support, and epidemiological relevance (9). The Nextstrain, which is a web-based platform for visualizing pathogen evolution in real time (10), has its own clade naming system called Nextstrain clade. Nextstrain clade designates a new clade by taking into account the regional and global frequencies of viral sequences (11). However, both the Pango nomenclature system and the Nextstrain clade are not fully automated, and they require manual curation by experts or crowdsourcing. Global Initiative on Sharing All Avian Influenza Data (GISAID), which is a database system facilitating the rapid sharing of nucleotide sequences of viruses, has its own clade classification system for SARS-CoV-2. GISAID uses predefined marker mutations to designate a new clade (12). However, it is difficult to predefine marker mutations for viruses that emerge in the future in advance.

A few studies have proposed methods to automate the designation of a new lineage or clade using statistics on nucleotide sequences or the phylogenetic tree. McBroom et al. proposed an automated lineage designation system, called Autolin, aiming to fully automate the Pango system (13). Autolin designates a new lineage using a metric called the genotype representation index (GRI). For a given focal ancestral node, the GRI of a node *v* is defined as the ratio between the branch length from the focal ancestral node to *v* and the total of branch lengths from *v* to tip nodes descending from *v*. VanderWaal et al. introduced a phylogenetic-based classification method for porcine reproductive and respiratory syndrome virus-type 2 sequences, comparing the robustness and reproducibility of 140 approaches for fine-scale classification of ORF5 sequences (14). Methods proposed by these two studies relied purely on phylogenetic information and do not consider phenotypic aspects of viruses. Chiara et al. developed a framework called HaploCoV using the unsupervised classification technique (15). HaploCoV classifies SARS-CoV-2 sequences into haplogroups, which are groups of viral sequences sharing specific combinations of mutations. HaploCoV also has a functionality to detect emerging variants, which is achieved by monitoring frequencies of haplogroups through time and space, and/or by using the VOC-ness score calculated from the genetic information. The VOC-ness score is used to associate the phenotypic information of viruses with the emerging haplogroups, but it is difficult to determine the phenotype of emerging haplogroups only from functionally annotated mutations. Hence, there is no existing nomenclature system that can automatically designate a new lineage or clade by considering both genetic and phenotypic similarity among virus strains.

Transmissibility, host range, and pathogenicity are important phenotypes to define variants of a virus. Among these phenotypes, transmissibility, which consists of intrinsic transmissibility and immune escaping ability, has been used to define variants of SARS-CoV-2 (2). The transmissibility of an infectious disease in an ongoing epidemic can be quantified using the effective reproduction number (*R*_*t*_), which is the average number of secondary cases generated by a primary case at time *t* (16). However, it is not straightforward to use *R*_*t*_ to quantify the transmissibility of a variant for two reasons. First, multiple variants can be circulating in the same population simultaneously. To calculate the *R*_*t*_ of each variant, the total number of new cases at time *t* needs to be decomposed into the numbers of new cases of variants. This is not a critical problem because the number of new cases of a variant can be estimated by multiplying the total number of new cases and the relative frequency of the variant estimated from a sampled population using viral genome sequencing.

Second, *R*_*t*_ of a variant can change over time not only from viral factors, but also from non-viral factors such as host behaviour, herd immunity, temperature, and humidity. One approach to separate viral factors from such time-dependent quantities is to calculate the ratio of *R*_*t*_ of a variant relative to *R*_*t*_ of another variant. This ratio is called the relative effective reproduction number. Several studies have estimated the relative effective reproduction number among SARS-CoV-2 variants based on temporal changes in observed frequencies of SARS-CoV-2 variants during original Wuhan–Alpha replacement (17), Alpha–Delta replacement (18, 19), Delta–Omicron replacement (20–22). The relative effective reproduction numbers among Omicron sub-lineages have also been investigated. Examples include BA.2 (23, 24), BA.4 and BA.5 (25), BA.2.75 (26), XBB.1.5 (27), XBB.1.6 (28), BA.2.86 (29), EG.5.1 (30), HK.3 (31), JN.1 (32), KP.2 (33), KP.3, LB.1 and KP.2.3 (34), KP.3.1.1 (35), XEC (36), and LP.8.1 (7). Most of these studies calculated the relative effective reproduction number among the lineages using the counts of sequences assigned to Pango lineages, and Pango nomenclature system is essential to calculate the relative effective reproduction number among circulating viruses.

Since the emergence of Omicron, the Pango nomenclature system has designated more than 1,700 lineages as Omicron subvariants and Omicron recombinants (37). Given that a large number of variants are circulating in the same population at the same time, it is difficult to interpret the evolutionary dynamics of Omicron sub-lineages. For instance, Kaku et al. calculated the relative effective reproduction numbers of 140 Omicron sub-variants compared to EG.5.1 in the United Kingdom (32). Looking at their results carefully, one can notice that multiple Pango lineages share almost the same value of relative effective reproduction numbers. For instance, 21 Pango lineages have the relative effective reproduction numbers of 1.00±0.01 in the United Kingdom (from EG.5.3 to GK.2 in pp.12 in Appendix of the paper by Kaku et al.). This means that these 21 lineages had almost the same transmissibility as EG.5.1 in the United Kingdom. Moreover, these 21 lineages included five immediate sub-lineages of EG.5.1 (i.e., EG.5.1.1, EG.5.1.3, EG.5.1.4, EG.5.1.5, EG.5.1.7). These data suggest that the Pango nomenclature system may have designated a group of viruses into more than one lineage even if they share the same transmissibility and similar genetic background. There is a possibility that we can construct a simpler model to describe lineage dynamics by assuming that multiple Pango lineages can have the same transmissibility. However, there is no existing method to estimate relative effective reproduction numbers among viral lineages by allowing multiple lineages to share the same relative effective reproduction numbers.

Let us consider a model allowing multiple lineages to share the same parameter for their relative effective reproduction numbers. Then, such a model can have fewer number of free parameters than the original model in which all lineages have different parameters of the relative effective reproduction numbers. According to the principle of Occam’s razor, of two competing theories, the simpler explanation of an entity is to be preferred (38). In other words, models should be selected by considering a good balance between the number of parameters and goodness of fit. One of the most popular implementations of the Occam’s razor principle is the Akaike Information Criterion (AIC), which evaluates models using the maximum likelihood and the number of parameters of the models (39). Using this criterion, one can consider a model to be a better model than another model when the former has a lower AIC value than the latter.

To model the lineage dynamics where multiple lineages may share the same relative effective reproduction numbers, we need to determine for each pair of lineages whether the two lineages share the same relative effective reproduction number or not. In other words, we need to partition a set of lineages into several groups of lineages that share the same relative effective reproduction numbers. This problem is mathematically equivalent to the set partitioning problem (40). Suppose we have *n* lineages of a virus. If we assume that two out of *n* lineages have the same relative effective reproduction numbers, then we have *n*(*n* − 1)/2 choices of such lineages and *n* lineages have *n* − 1 different relative reproduction numbers. Similarly, we also need to consider the cases where *n* lineages have *n* − 2, *n* − 3, …, different relative effective reproduction numbers. It is known that the number of partitions of *n* objects is equal to the (*n* + 1)th Bell number (41). When we try to find the lineage partitions that has the lowest AIC value among all possible lineage partitions, a naive approach is to enumerate all possible partitions. However, this approach quickly becomes impractical when we analyse a large number of lineages, because the Bell number grows faster than the exponential functions of *n*. Therefore, it is necessary to develop a method that can efficiently find the best lineage partition among all possible lineage partitions.

In this paper, we propose a new algorithm, called FindPart-*w*, which can identify groups of viral lineages that share the same relative effective reproduction numbers. To this end, we extend the previous model of lineage replacement by Ito et al. (18) to estimate relative effective reproduction numbers among lineages under constraints where multiple lineages may share the same relative effective reproduction numbers. We evaluate the computational time and the accuracy of the FindPart-*w* algorithm in identifying viral lineages sharing the same reproduction numbers using hypothetical observation datasets created by simulations. Finally, we apply the FindPart-*w* to real-world data from the SARS-CoV-2 epidemic in the United States, exploring which circulating lineages could have the same relative effective reproduction numbers.

## Materials and Methods

### The generation time of SARS-CoV-2 Omicron sub-variants

The generation time is the interval between the infection of an individual of a primary case and the infection of a secondary case (42). We assumed that the generation time of infections by Omicron sub-variants followed a gamma distribution with a shape parameter of *α* = 4.11 and a scale parameter of *θ* = 0.73. These parameters were calculated so that the gamma distribution had a mean of *μ* = 3.0 and a variance of *σ*^2^ = 2.15 (43). We truncated the generation time distributions at *t* = 1 and *t* = *l* and discretized by 1 day as follows:

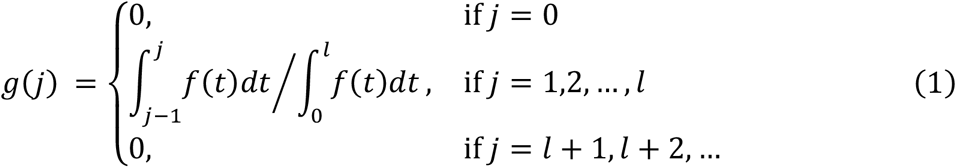

where *f*(*t*) is the probability density function of the gamma-distributed generation. We set *l* = 4, which is the 99 percent quantile of *f*(*t*).

### Lineage Replacement Model with Relative Instantaneous Reproduction Number

There are two ways to define *R*_*t*_, the case reproduction number and the instantaneous effective reproduction number. The case reproduction number, *R*_*c*_(*t*), is defined as the average number of people an individual infected at time *t* can expect to infect, while the instantaneous effective reproduction number, *R*(*t*), is defined as the average number of people an individual infected at time *t* could expect to infect if conditions remained unchanged (44). In this study, we used *R*(*t*) as the definition of *R*_*t*_, since the *R*(*t*) can be estimated using case data up to *t*, and it is more appropriate for the real-time analysis of an epidemic than *R*_*c*_(*t*).

The ratio of *R*(*t*) of a lineage relative to *R*(*t*) of another variant is called the relative instantaneous reproduction number (*R*_*RI*_), and Ito et. al. proposed a mathematical model of lineage replacement using *R*_*RI*_ (18). In this paper, we refer to this model as the original RelRe model. Here, we summarize the original RelRe model. We use different names and indices of variables from Ito et al.’s paper, in order to clearly describe how we will extend the original RelRe model to allow multiple lineages to share the same *R*_*RI*_.

Let *V* = {*v*_0_, *v*_1_, …, *v*_*n*−1_} be a set of *n* viral lineages that are circulating in a population. We refer to *v*_0_ as the baseline lineage, and we calculate the *R*_*RI*_ of non-baseline lineages *v*_1_, …, *v*_*n*−1_ with respect to (w.r.t.) *v*_0_. The non-baseline lineages *v*_1_, …, *v*_*n*−1_ are called subject lineages.

Suppose 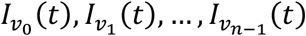 are the numbers of new infections by *v*_0_, …, *v*_*n*−1_ at time *t*, respectively. Then, the total number of new infections at time *t, I*(*t*), is represented by

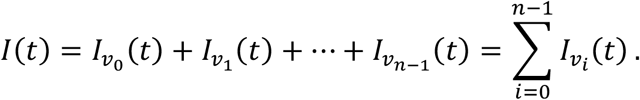

Let 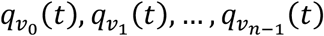 be the relative frequencies of lineages *v*_0_, *v*_1_ …, *v*_*n*−1_ at time *t*, respectively. The relative frequency of lineage *v*_*i*_ at time *t* can be represented by

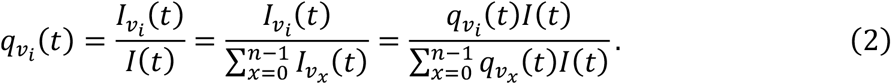

By the definition by Fraser et. al. (44), the *R*(*t*) of infections by lineages *v*_*i*_ at *t* is given as follows:

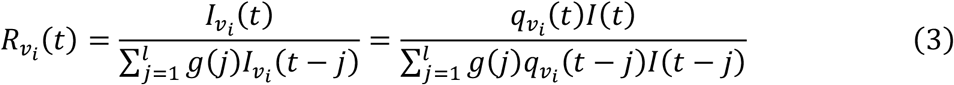

for *i* = 0, 1, …, *n* − 1, where *g*(*j*) is a probability mass function of the generation time defined in Equation (1). We assume that the generation time distribution is the same for all lineages. Transforming Equation (3), we get

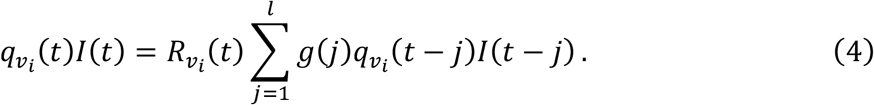

Substituting Equation (4) to Equation (2), we get

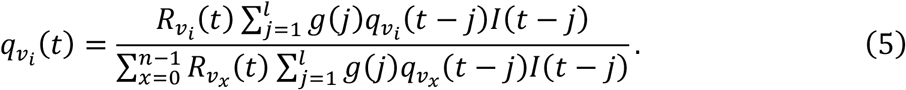

We assume that 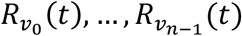 are *ρ*_0_, …, *ρ*_*n*−1_ times larger than that of the baseline lineage *v*_0_ at *t* for any *t*, i.e.,

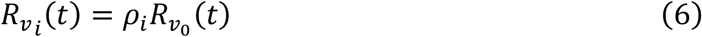

for *i* = 0, 1, …, *n* − 1. The constant *ρ*_*i*_ is the relative instantaneous reproduction number, *R*_*RI*_, of *v*_*i*_ w.r.t. *v*_0_. Note that *ρ*_0_, *R*_*RI*_ of *v*_0_ w.r.t. *v*_0_, is always 1.0 by definition. Rewriting Equation (5) using Equation (6), and dividing the numerator and denominator of the right-hand side by 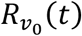, we get

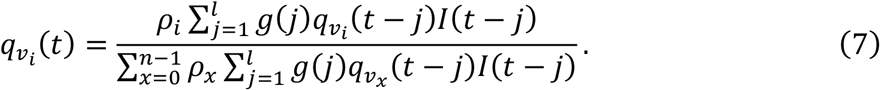

We also assume that the following approximation holds for each *j* = 1,2, …, *l* for any *t*:

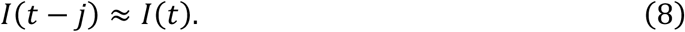

Then, Equation (7) can be simplified as follows:

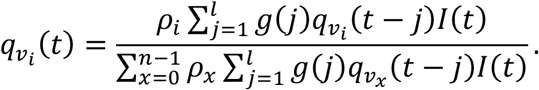

Dividing the numerator and the denominator of the right-hand side by *I*(*t*), we get

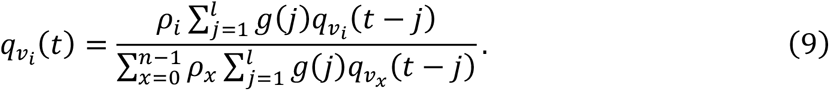

Since 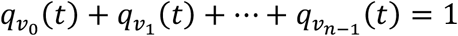, we use Equation (9) for subject lineages *v*_*i*_ for *i* = 1,2, …, *n* − 1 and use

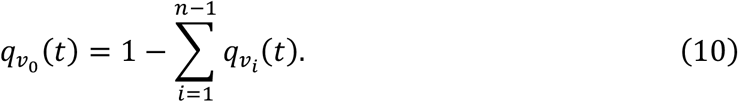

for the baseline lineage *v*_0_.

### Calculation of the Trajectory of Lineage Replacement

Let *t*_*s*_ and *t*_*e*_ be the calendar times at the start and end of the target period of analysis, respectively. We suppose all lineages *v*_0_, …, *v*_*n*−1_ existed at *t*_*s*_. Then 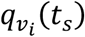 is considered as a parameter that denotes the relative frequency of a subject lineage *v*_*i*_ in the population at *t*_*s*_, where *i* = 1,2, …, *n* − 1. From Equation (10), the relative frequency of the baseline lineage *v*_0_ at *t*_*s*_ is given by

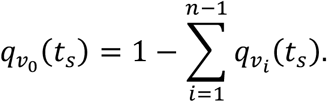

The relative frequencies of subject lineages *v*_1_, …, *v*_*n*−1_ are determined by recursively applying Equation (9) for *t* = *t*_*s*_ + 1, *t*_*s*_ + 2, …. The calculation of 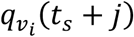 using Equation (9) requires values of 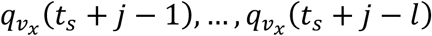 for *x* = 1, …, *n* − 1. Thus, relative frequencies of lineages before *t*_*s*_ are required to calculate the relative frequencies of lineages for the first *l* − 1 days after *t*_s_. We use the value of 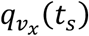 for 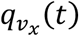 for *t* < *t*_*s*_ for *x* = 1, …, *n* − 1. The effect of this approximation is negligible if we assume the duration between *t*_*s*_ and *t*_*e*_ is much longer than *l*.

### Likelihood Function

Let 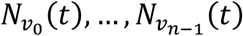 be numbers of lineages *v*_0_, …, *v*_*n*−1_ observed at a calendar time *t* where *t* = *t*_*s*_, *t*_*s*_ + 1, …, *t*_*e*_. The likelihood function of parameters *ρ*_1_, …, *ρ*_*n*−1_ and 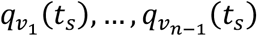 under the RelRe model for observing *v*_0_, …, *v*_*n*−1_ respectively 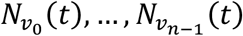 times at *t* is given by the formula:

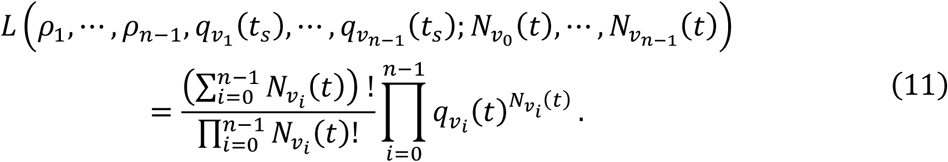

Multiplying the likelihood at *t* = *t*_*s*_, *t*_*s*_ + 1, …, *t*_*e*_, we obtain the likelihood of the parameters under the RelRe model for the entire period as follows:

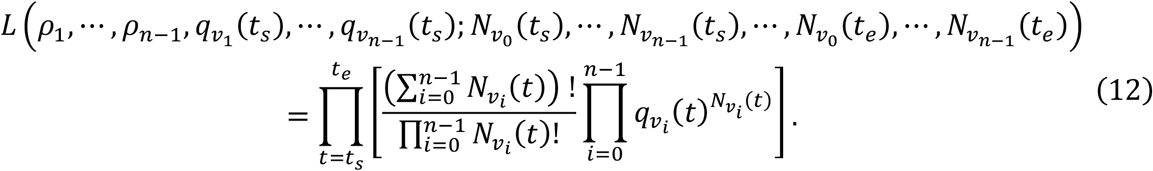

### Partition of Lineages and Constrained RelRe Model

The original RelRe model by Ito et al. (18) assumes that lineages *v*_0_, …, *v*_*n*−1_ have different *R*_*RI*_ from each other. In other words, *ρ*_0_, …, *ρ*_*n*−1_ are assumed to have different values. This is also true for other lineage replacement models with relative effective reproduction numbers, such as one used by Suzuki et al. (22). When we have *n* lineages, the likelihood function in Equation (12) has 2*n* − 2 free parameters, consisting of *n* − 1 free parameters for *R*_*RI*_ and *n* − 1 free parameters for the initial frequencies of subject lineages. In this paper, we extend the original RelRe model by introducing equality constraints among *R*_*RI*_ of lineages, *ρ*_0_, …, *ρ*_*n*−1_. Introduced equality constraints contribute to reduce the number of free parameters of the RelRe model. For instance, if we introduce an equality constraint *ρ*_*i*_ = *ρ*_*j*_ for a pair of *i* and *j* such that *i* ≠ *j*, the number of free parameters in this constrained model becomes 2*n* − 3. Meanwhile, a large number of constrained models can be defined by introducing equality constraints among *ρ*_0_, …, *ρ*_*n*−1_. To discuss the number of such models, we formulate this problem using the set partitioning problem (40).

#### Definition 1

(A partition of lineages) Let *V* = {*v*_0_, *v*_1_, …, *v*_*n*−1_} be a set of lineages. A set of lineage sets *P* = {*B*_0_, *B*_1_, …, *B*_*h*−1_} is called a partition of *V* if and only if 1) none of *B*_*j*_ ∈ *P* is empty, 2) *B*_0_ ∪ *B*_1_ ∪ … ∪ *B*_*h*−1_ = *V*, 3) *B*_*i*_ ∩ *B*_*j*_ is empty for any *B*_*i*_, *B*_*j*_ ∈ *P* if *i* ≠ *j*. Each element of a partition, *B*_*i*_ ∈ *P*, is called a block. The number of blocks, *h*, is called the rank of *P*.

**Example 1** Let *V* = {*v*_0_, *v*_1_, *v*_2_} be the set of lineages. There are five partitions of *V*: *P*_1_ = {{*v*_0_}, {*v*_1_}, {*v*_2_}}, *P*_2_ = {{*v*_0_, *v*_1_}, {*v*_2_}}, *P*_3_ = {{*v*_0_, *v*_3_}, {*v*_2_}}, *P*_4_ = {{*v*_0_}, {*v*_1_, *v*_2_}}, *P*_5_ = {{*v*_0_, *v*_1_, *v*_2_}}. These partitions are all the partitions of *V* and there is no other partition of *V* than these. The rank of *P*_1_, *P*_2_, *P*_3_, *P*_4_, and *P*_5_ are 3, 2, 2, 2, and 1, respectively.

For a given set of *n* lineages, each partition of the set can be represented by an integer vector of length *n* called a restricted growth string (40).

#### Definition 2

(Restricted growth string) A vector of non-negative integers (*b*_0_, *b*_1_, …, *b*_*n*−1_) is called a restricted growth string (RGS) if and only if

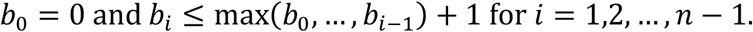

For each partition *P* of *V*, there exists exactly one RGS representing *P* (45). An RGS *X* = (*b*_0_, *b*_1_, …, *b*_*n*−1_) represents a partition *P* such that if *b*_*i*_ = *b*_*j*_ then *v*_*i*_ and *v*_*j*_ belong to the same block in *P* for any pair of *i* and *j*. The integer *b*_*n*−1_ is equal to *h* − 1, where *h* is the rank of *P*. The condition of restricted growth of integers in Definition 2 ensures the uniqueness of the RGS representation for each partition.

**Example 2** (Continued from Example 1) the RGS corresponding to *P*_1_, *P*_2_, *P*_3_, *P*_4_, and *P*_5_ in Example 1 are

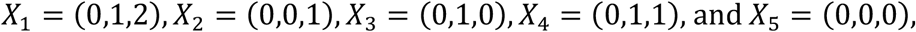

respectively.

When *X* is an RGS notation of a partition *P, P* is denoted by *P*_*X*_.

#### Definition 3

(Refinement and aggregation) Suppose *P* and *P*′ are partitions of *V. P*′ is called a refinement of *P*, if for every block 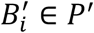 there exists a block *B*_*j*_ ∈ *P* such that 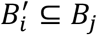. Partition *P*′ is called an immediate refinement of *P* if the rank of *P* is *h* and the rank of *P*′ is *h* + 1. Partition *P* is called an aggregation of partition *P*′ if *P*′ is a refinement of *P*. Partition *P* is called an immediate aggregation of *P*′ if *P*′ is an immediate refinement of *P*. Note that any partition is both a refinement and an aggregation of itself.

Partitions of lineages can be used to reduce number of the parameters of a model by introducing equality constraints among the parameters of lineages. Now we consider applying this idea to the lineage replacement model.

#### Definition 4

(Constrained RelRe model) Let *V* = {*v*_0_, *v*_1_, …, *v*_*n*−1_} be a set of lineages and *P* = {*B*_0_, …, *B*_*h*−1_} be a rank-*h* partition of *V*. Suppose *R*_*RI*_ of *v*_0_, …, *v*_*n*−1_ w.r.t *v*_0_ are *ρ*_0_, …, *ρ*_*n*−1_, respectively. Then, *M*_*P*_ represents a RelRe model that has a constraint *ρ*_*i*_ = *ρ*_*j*_ if and only if *v*_*i*_ and *v*_*j*_ belong to the same block *B*_*u*_ ∈ *P*. We call *M*_*P*_ as the RelRe model constrained by *P*, and we call *P* as the constraining partition of *M*_*P*_. If *X* is an RGS notation of *P*, then the model *M*_*P*_ is also denoted by *M*_*X*_.

From the definition of *R*_*RI*_, *ρ*_0_ is always equal to one. So *ρ*_0_ is not a free parameter. A RelRe model constrained by a rank-*h* partition has *h* − 1 free parameters for *R*_*RI*_ and *n* − 1 free parameters for initial frequencies and thus a total of *h* + *n* − 2 free parameters. There may be a lineage *v*_*i*_ such that *ρ*_*i*_ is equal to *ρ*_0_. Such *v*_*i*_ needs to belong to the same block as *v*_0_. For this reason, we need to include *v*_0_ in a block in the partition of *V*, even if *ρ*_0_ is always equal to one.

#### Definition 5

(The finest model and the coarsest model) Consider a set of *n* lineages *V* = {*v*_0_, …, *v*_*n*−1_}. The partition

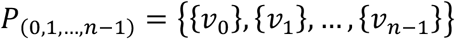

is called the finest partition, which is a refinement of any partition of *V*. The RelRe model constrained by *P*_(0,1,…,*n*−1)_ is called the finest model of *V*, and it is denoted by *M*_(0,1,…,*n*−1)_. The partition

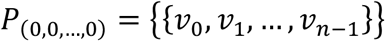

is called the coarsest partition, which is an aggregation of any partition of *V*. The RelRe model constrained by *P*_(0,0,…,0)_ is called the coarsest model of *V*, and it is denoted by *M*_(0,0,…,0)_.

The original RelRe model, which has no equality constraints among parameters *ρ*_0_, …, *ρ*_*n*−1_ is equivalent to the finest model *M*_(0,1,…,*n*−1)_. In contrast, all *ρ*_0_, …, *ρ*_*n*−1_ in the coarsest model *M*_(0,0,…,0)_ are constrained to be one, since *ρ*_0_ is always one.

**Example 3** Suppose we have four lineages *V* = {*v*_0_, *v*_1_, *v*_2_, *v*_3_}. Let *ρ*_0_, *ρ*_1_, *ρ*_2_, and *ρ*_3_ be the *R*_*RI*_ of *v*_0_, *v*_1_, *v*_2_, and *v*_3_ w.r.t *v*_0_, respectively. The number of possible partitions of *V* is 15. This means that there exist 15 constrained RelRe models. Figure 1 shows a partition lattice consisting of all 15 partitions of *V. P*_(0,1,2,3)_ at the top is the finest partition

**Figure 1.**
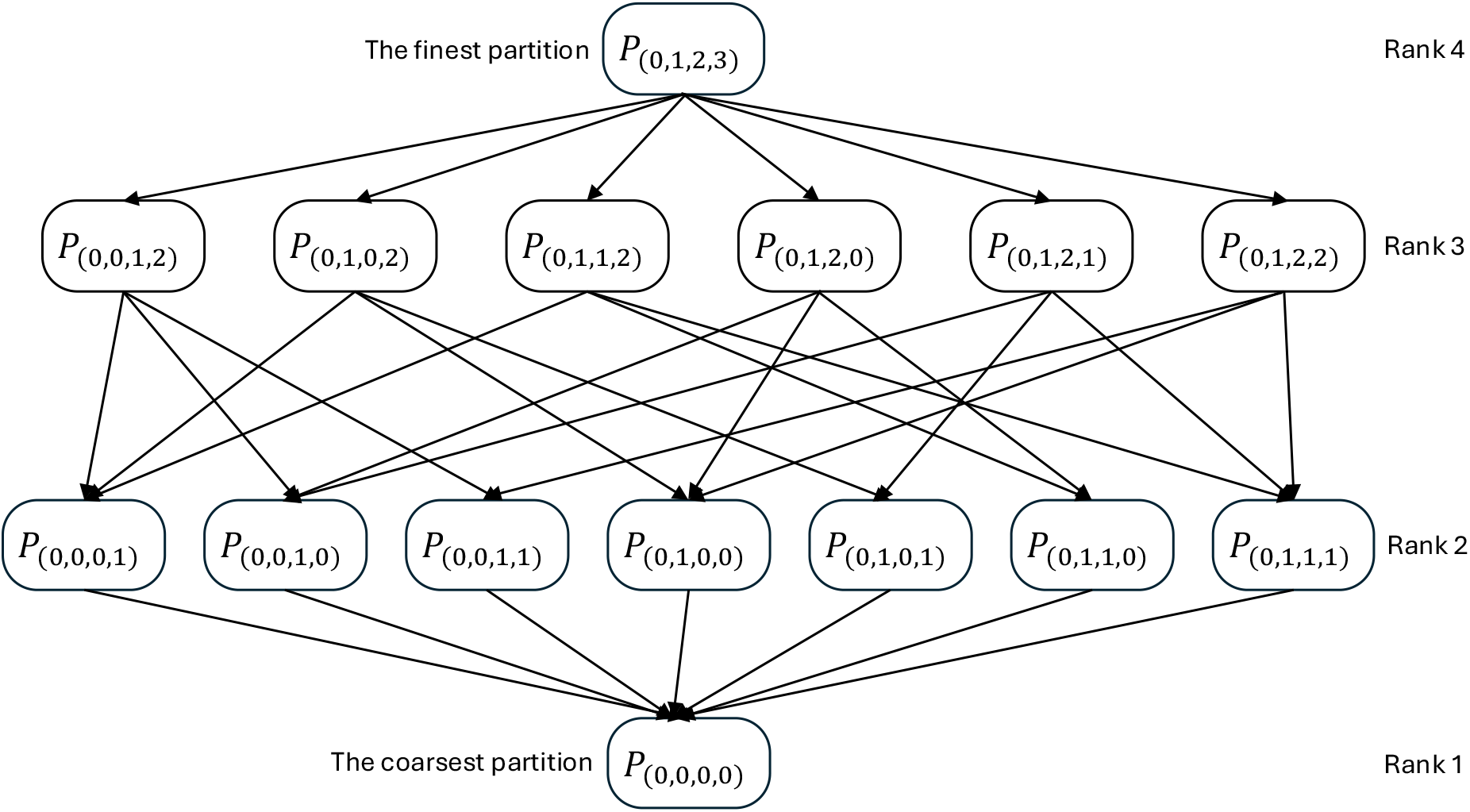
The partition lattice of four lineages. Each rounded rectangle represents a unique partition. Each arrow indicates a refinement-aggregation relationship between two partitions. A partition at which an arrow ends is an aggregation of the partition at which the arrow starts. A partition at which an arrow starts is a refinement of the partition at which the arrow ends. Partitions that are horizontally aligned in the lattice have the same rank. The finest partition on the top has a rank of four, while the coarsest partition at the bottom has a rank of one.

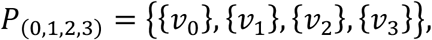

and *ρ*_0_, *ρ*_1_, *ρ*_2_, and *ρ*_3_ in its corresponding model, *M*_(0,1,2,3)_, can have different values. The partition *P*_(0,0,0,0)_ at the bottom is the coarsest partition

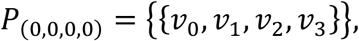

and *ρ*_0_, *ρ*_1_, *ρ*_2_, and *ρ*_3_ in its corresponding model, *M*_(0,0,0,0)_, are constrained to be the same value, one. An arrow from one partition in rank *h* to another in rank *h* − 1 indicates that the lower one is an immediate aggregation of the upper one, and the model corresponding to the lower partition is obtained by introducing a new constraint into the model corresponding to the upper partition. For example, if we introduce a new constraint *ρ*_0_ = *ρ*_1_ into *M*_(0,1,2,3)_ so that lineages *v*_0_ and *v*_1_ share the same *R*_*RI*_, then we obtain *M*_(0,0,1,2)_, which is the RelRe model constrained by

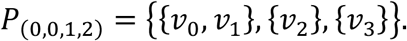

Similarly, if we introduce another constraint *ρ*_2_ = *ρ*_3_ into *M*_(0,0,1,2)_ so that lineages *v*_2_ and *v*_3_ share the same *R*_*RI*_, then we obtain *M*_(0,0,1,1)_, which is the RelRe model constrained by

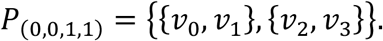

Finally, if we introduce another constraint *ρ*_1_ = *ρ*_2_ into *M*_(0,0,1,1)_ so that lineages *v*_1_ and *v*_2_ share the same *R*_*RI*_, then we obtain *M*_(0,0,0,0)_, which is the coarsest model.

The following proposition on the number of partitions is known (41).

#### Proposition 1

For a given set *V* of size *n*, the number of partitions of *V* is equal to the (*n* + 1)th Bell number.

From Proposition 1, one can know that, for instance, there exist 15 partitions when *n* = 4, 115,975 partitions when *n* = 10, and 158,235,372 partitions when *n* = 20.

## Algorithm to Find the Best Constrained RelRe Model

### Akaike Information Criterion

Now, we consider the problem of finding a good constrained RelRe model for a given dataset. We use the Akaike Information Criterion (AIC) to measure the goodness of a model (39). The AIC is defined as follows:

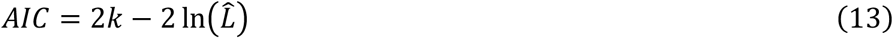

where 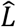 is the maximum likelihood of a model with *k* free parameters.

Consider a set of lineages *V* = {*v*_0_, …, *v*_*n*−1_}, where *v*_0_ is the baseline lineage. The original RelRe model, which has 2*n* − 2 free parameters, may not always be the best model in terms of AIC. The reason for this is that some lineages may have the same transmissibility even though they have different nucleotides on their genome. If this is the case, a RelRe model having equality constraints *ρ*_*i*_ = *ρ*_*j*_ for such lineages may have a smaller AIC, as the introduced equality constraints reduce the number of free parameters.

### Maximum Likelihood Estimation

Let *V* = {*v*_0_, …, *v*_*n*−1_} be a set of viral lineages circulating in the population, *v*_0_ be the baseline lineage, and *v*_1_, …, *v*_*n*−1_ be subject lineages. From the observed counts of these lineages, we estimated parameters 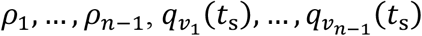 under the original RelRe model without any equality constraints among *ρ*_0_, …, *ρ*_*n*−1_. This estimation was conducted by maximizing the likelihood defined in Equation (12). Initial values for *ρ*_1_, …, *ρ*_*n*−1_ were set to 1.0, and those for 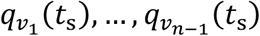 were set to 0.001. We then calculated the AIC for the original RelRe model according to Equation (13), setting In 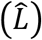 to the logarithm of the maximum loglikelihood and *k* to 2*n* − 2, which was the number of free parameters in the RelRe model. This gives the AIC of the finest model. The 95% confidence interval (C.I.) of each parameter was calculated using the profile likelihood method (46).

Suppose a rank-*h* partition *P* = {*B*_0_, …, *B*_*h*−1_} of *V* was given to specify equality constraints among *ρ*_0_, …, *ρ*_*n*−1_. We performed maximum likelihood estimation for *M*_*P*_, the RelRe model constrained by *P*, by maximizing the likelihood defined in Equation (12) subject to *ρ*_*i*_ = *ρ*_*j*_ for all (*i, j*) such that *v*_*i*_ and *v*_*j*_ belong to the same block *Bu* ∈ *P*. Block *B*_0_ is assumed to contain *v*_0_, and for each lineage *v*_*i*_ in *B*_0_, *ρ*_*i*_ was fixed to 1.0 and excluded from free parameters. For each 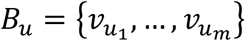 where *u* = 1,2, …, *h* − 1, the values of 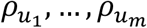 were represented by a single representative parameter 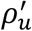. The maximum likelihood estimation was performed by maximizing the likelihood defined by Equation (12) by varying *h* + *n* − 2 free parameters, 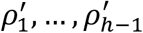 and 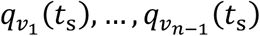.

We reused the maximum likelihood estimates of the finest model as initial values of maximum likelihood estimation of the other constrained RelRe models. Let 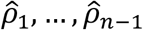 be the maximum likelihood estimates of the *R*_*RI*_ of *v*_1_, …, *v*_*n*−1_ and 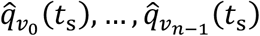 be that of the initial frequencies estimated under the finest model. For each block 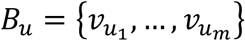 in *P* where *u* = 1,2, …, *h* − 1, we set the initial value of 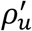 to the mean of 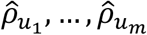. The initial values of 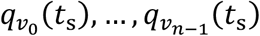 were set to 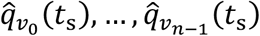, respectively.

### Properties of AIC of Constrained RelRe Models

Finding the constrained RelRe model that has the smallest AIC among all constrained RelRe models is not a trivial matter. A naïve approach is to calculate the AIC of all constrained RelRe models by exhaustively enumerating their constraining partitions using Hutchinson’s algorithm (45). Although the naïve approach always finds the constrained RelRe model having the smallest AIC, the method is impractical, since the number of possible partitions for *n* lineages grows faster than exponential functions.

Let *V* = {*v*_0_, …, *v*_*n*−1_} be a set of lineages and *P*_*X*_ and *P*_*Y*_ be partitions of *V*. Suppose *M*_*X*_ and *M*_*Y*_ are RelRe models constrained by *P*_*X*_ and *P*_*Y*_, respectively. The following lemma gives the relationship between the maximum likelihood values of *M*_*X*_ and *M*_*Y*_ when *P*_*X*_ is a refinement of *P*_*Y*_.

#### Lemma 1

Let *P*_*X*_ and *P*_*Y*_ be partitions of *V*. If *P*_*X*_ is a refinement of *P*_*Y*_, then the maximum likelihood of *M*_*X*_ is greater than or equal to the maximum likelihood of *M*_*Y*_.

**Proof** Let 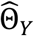 be the parameters such that 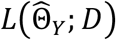 is the maximum likelihood of *M*_*Y*_ for a given dataset *D*. Since *P*_*Y*_ is an aggregation of *P*_*X*_, if *ρ*_*i*_ = *ρ*_*j*_ is a constraint in *M*_*X*_ then *ρ*_*i*_ = *ρ*_*j*_ is also a constraint in *M*_*Y*_. It follows from its contraposition that if *ρ*_*i*_ = *ρ*_*j*_ is not a constraint in *M*_*Y*_ then *ρ*_*i*_ = *ρ*_*j*_ is not a constraint in *M*_*X*_. Thus, 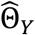is also within the parameter space of *M*_*X*_. Therefore, the maximum likelihood of *M*_*X*_ is greater than or equal to 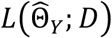, which is the maximum likelihood of *M*_*Y*_. □

The following lemma gives the relationship between the AIC values of *M*_*X*_ and *M*_*Y*_ when *P*_*X*_ is a refinement of *P*_*Y*_.

#### Lemma 2

Let *P*_*X*_ and *P*_*Y*_ be partitions of *V* having a rank of *h*_*X*_ and *h*_*Y*_, respectively. If *P*_*X*_ is a refinement of *P*_*Y*_, then

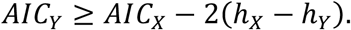

where *AIC*_*Y*_ and *AIC*_*X*_ are the AIC of *M*_*Y*_ and that of *M*_*X*_, respectively.

**Proof**. The numbers of free parameters in *M*_*X*_ and *M*_*Y*_ are *h*_*X*_ + *n* − 2 and *h*_*Y*_ + *n* − 2, respectively. Here, *h*_*X*_ ≥ *h*_*Y*_, because *h*_*X*_ is a refinement of *h*_*Y*_. From the definition of AIC, it follows that

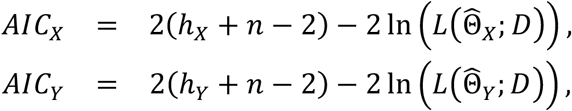

where 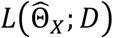 and 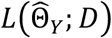 are the maximum likelihood of *M*_*X*_ and *M*_*Y*_ for given data *D*. From Lemma 1

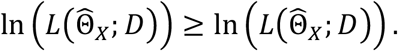

From these equations, we get

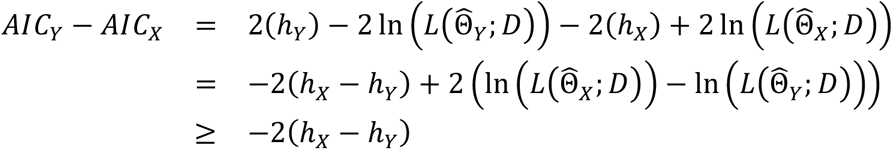

Therefore, it follows that

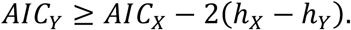

#### Lemma 3

Suppose partition *P*_*X*_ is a refinement of partition *P*_*Y*_ and the rank of *P*_*X*_ is *h*_*X*_, then the *AIC*_*Y*_ ≥ *AIC*_*X*_ − 20*h*_*X*_ − 1).

**Proof** From Lemma 2 it holds that *AIC*_*Y*_ ≥ *AIC*_*X*_ − 2*h*_*X*_ + 2*h*_*Y*_, where *h*_*Y*_ is the rank of *P*_*Y*_. Because the smallest value of *h*_*Y*_ is 1, *AIC*_*Y*_ ≥ *AIC*_*X*_ − 2*h*_*X*_ + 2. □

The value of *AIC*_*X*_ − 2(*h*_*X*_ − 1) gives the lower bound of AIC for all RelRe models constrained by an aggregation of *P*_*X*_. Our objective is to find the constrained RelRe model that has the smallest AIC. Suppose we have calculated *AIC*_*X*_ and *AIC*_*X*_ for partitions *P*_*X*_ and *P*_*Z*_. If *AIC*_*Z*_ − 2(*h*_*X*_ − 1) > *AIC*_*Z*_ and *P*_*Y*_ is an aggregation of *P*_*X*_, then it holds that *AIC*_*Y*_ ≥ *AIC*_*X*_ − 2(*h*_*X*_ − 1) > *AIC*_*Z*_ from Lemma 3. We can know that any RelRe model constrained by an aggregation of *P*_*X*_ has a larger AIC than the RelRe model constrained by *P*_*Z*_ without computing their maximum likelihood. We can use this property to reduce the number of maximum likelihood estimations when searching for the constrained RelRe model that has the minimum AIC.

### Algorithm FindPart-*w*

Given that a naïve approach is impractical for larger sizes of *n*, we give an algorithm, called FindPart-*w*, that utilizes Lemma 3 to reduce the number of maximum likelihood estimations in searching for the constrained RelRe model having the smallest AIC (Figure 2). The algorithm starts with the maximum likelihood estimation using the finest model of which rank *h* = *n*, which is the original RelRe model. Then, the algorithm iterates over *h* from *n* − 1 to 1, processing partitions with rank *h*. The integer *w* ranges from 1 to ∞, and it represents the maximum number of partitions that are passed to the next rank when the search at a rank was finished.

**Figure 2.**
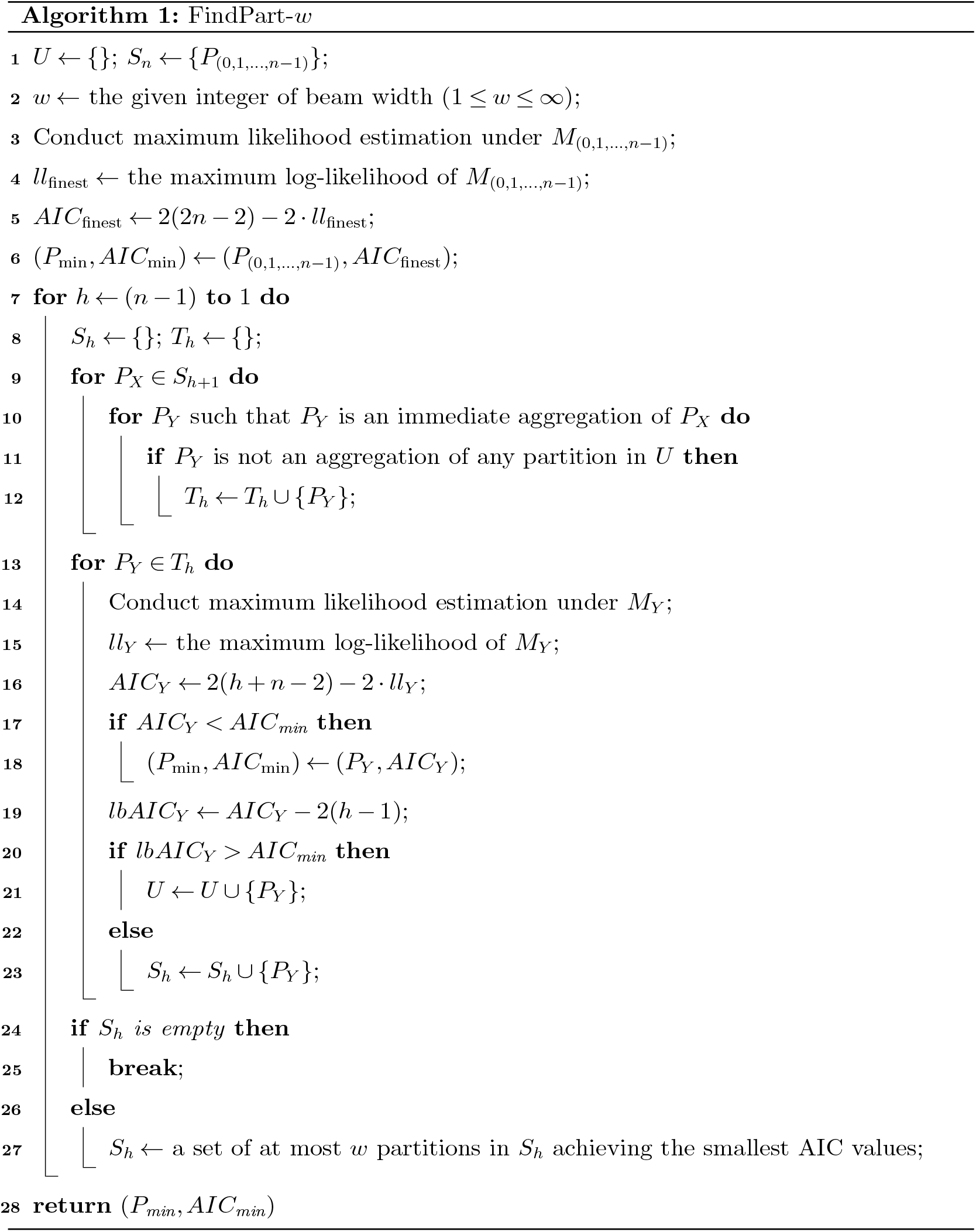
The FindPart-*w* algorithm.

The algorithm uses sets of partitions, *U* and *S*_*n*_, *S*_*n*−1_, …, *S*_1_. The set *U* stores partitions of which aggregations have no chance of being the constraining partition of the constrained RelRe model having the minimum AIC. The set *U* is initialized to an empty set at the beginning of the algorithm (line 1). Every partition added to *U* remains in *U* until the algorithm ends, *U* may contain partitions with different ranks. The set *S*_*h*_ where *h* = *n, n* − 1, …, 1 stores *h*-rank partitions of which aggregations have a chance of being the constraining partition of the constrained RelRe model having the minimum AIC. *S*_*n*_ is initialized to a set containing only the finest partition at the beginning of the algorithm (line 1). The maximum width of the beam *w* can be any positive integer including ∞, and the user can specify the value of *w* (line 2).

The algorithm starts with the evaluation of the finest partition, of which rank is *n*. The maximum likelihood estimation for the original RelRe model is conducted to determine the maximum log-likelihood and AIC of the finest model, *M*_(0,1,…,*n*−1)_ (lines 3, 4, and 5). While the algorithm is running, *P*_*min*_ and A*IC*_*min*_ respectively store the partition and AIC of the constrained RelRe model that has the minimum AIC among all constrained RelRe models that have been evaluated by the algorithm. Before the beginning of the main loop, the finest partition and the AIC of the finest model are recorded as *P*_*min*_ and as the *AIC*_*min*_, respectively (line 6).

The algorithm iterates evaluations of rank-*h* partitions, starting from *h* = *n* − 1 and decrementing to *h* = 1 (line 7). At the beginning of evaluations of rank-*h* partitions, *S*_*h*_ and the temporary partition set *T*_*h*_ are initialised as empty sets (line 8). At this point, any RelRe model constrained by an aggregation of a partition in *U* has no chance of having minimum AIC, while a RelRe model constrained by an aggregation of a partition in *S*_*h*+1_ has a chance of having minimum AIC among all partitions. For each partition *P*_*X*_ in *S*_*h*+1_ (line 9), the algorithm enumerates its immediate aggregation, *P*_*Y*_ (line 10), and checks whether if *P*_*Y*_ is an aggregation of a partition in *U* or not (line 11). Since any RelRe model constrained by an aggregation of a partition in *U* has no chance of having minimum AIC, *P*_*Y*_ is ignored if *P*_*Y*_ is an aggregation of a partition in *U*. Otherwise, *P*_*Y*_ is added to *T*_*h*_ if *P*_*Y*_ ∉ *T*_*h*_ (line 12).

For each partition *P*_*Y*_ in *T*_*h*_ (line 13), maximum likelihood estimation is conducted under the model *M*_*Y*_, of which equality constraints among *ρ*_0_, …, *ρ*_*n*−1_ are defined by *P*_*Y*_ (line 14). The value of the maximum log-likelihood of *M*_*Y*_ is stored as *ll*_*Y*_ (line 15). Then, the AIC of *M*_*Y*_, *AIC*_*Y*_, is calculated by using *ll*_*Y*_ (line 16). If *AIC*_*Y*_ is smaller than *AIC*_*min*_ (line 17), then *P*_*min*_ and A*IC*_*min*_ are updated to *P*_*Y*_ and *AIC*_*Y*_, respectively (line 18).

Then, *lbAIC*_*Y*_, which is the lower bound of AIC of all RelRe models constrained by an aggregation of *P*_*Y*_ is calculated based on Lemma 3 (line 19). If *lbAIC*_*Y*_ is greater than the minimum AIC up to this point, *AIC*_*min*_, (line 20), then *P*_*Y*_ is added to *U* (line 21). Otherwise, RelRe models constrained by some aggregations of *P*_*Y*_ have a chance of having the minimum AIC, thus *P*_*Y*_ is added to *S*_*h*_ (line 23).

At the end of each rank-*h* loop, we know that RelRe models constrained by an aggregation of the rank-*h* partition in *S*_*h*_ have a chance of having the minimum AIC. If *S*_*h*_ is empty (line 24), the algorithm breaks the main loop (line 25). Otherwise, if *w* ≠ ∞, the algorithm truncates *S*_*h*_ to include at most *w* partitions which achieve the smallest AIC values with their constrained RelRe models (line 27). Ideally, all partitions in *S*_*h*_ in the rank-*h* loop will be passed to and analysed in the rank-(*h* − 1) loop. To reduce the computational cost for searching, the algorithm takes a heuristic strategy. The heuristics assumes that an aggregation of a partition that achieves a small AIC is more likely to achieve a smaller AIC than an aggregation of a partition that achieves a large AIC. The algorithm does not truncate *S*_*h*_ and pass all partitions to the next rank if *w* = ∞.

The main loop of the algorithm continues until it finishes analysis of the coarsest partition, of which rank *h* = 1, or *S*_*h*_ becomes empty (line 25). The algorithm terminates by outputting the (*P*_*min*_, *AIC*_*min*_) (line 28).

#### Lemma 4.

Let 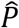 be a rank-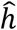 partition of *V* = {*v*_0_, …, *v*_*n*−1_} such that the RelRe model constrained by 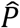 has the minimum AIC among all constrained RelRe models, and no other constrained RelRe model has the same AIC as 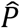. If the beam width, *w*, is set to ∞, then *S*_*h*_ contains at least one refinement of 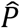 for all 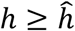 when FindPart-*w* terminates.

**Proof**. The Lemma can be proved by mathematical induction of rank for 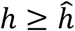. When *h* = *n*, the statement is true, because *S*_*n*_ contains the finest partition, *P*_(0,1,…,*n*−1)_, and *P*_(0,1,…,*n*−1)_ is a refinement of 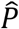.

Suppose the statement is true for *S*_*h*+1_, i.e., *S*_*h*+1_ contains the refinement of 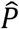. Let *P*_*X*_ in *S*_*h*−1_ be a refinement of 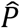. If 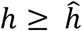, there exists a rank-*h* partition that is a refinement of 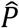 and an immediate aggregation of *P*_*X*_. Let 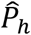 be such a rank-*h* partition. Note that 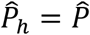 if 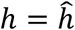. Since 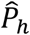 is an immediate aggregation of a partition in *S*_*h*+1_, the algorithm generates 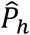 at line 10. From Lemma 3, any RelRe model constrained by an aggregation of a partition added to *U* has no chance of having minimum AIC (line 21). Therefore, for each partition *P*_*u*_ ∈ *U*, 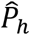 is not an aggregation of *P*_*u*_, because 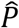, which has the minimum AIC among all constrained RelRe models, is an aggregation of 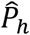. Thus, 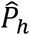 is added to *T*_*h*_ in line 12, and 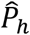 is evaluated in the loop starting from line 13. Since the AIC of the RelRe model constrained by 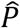 is smaller than or equal to *AIC*_*min*_, *lbAIC*_*Y*_ ≤ *AIC*_*min*_ when 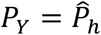. Thus, the condition *lbAIC*_*Y*_ > *AIC*_*min*_ in line 20 never becomes true for 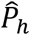, and 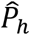 is added to *S*_*h*_ at line 23. Accordingly, *S*_*h*_ contains a refinement of 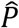, and the statement is true for rank *h*. By mathematical induction, we prove that the statement is true for all 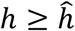. □

#### Theorem 1.

Let 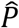 be a rank-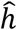 partition of *V* = {*v*_0_, …, *v*_*n*−1_} such that the RelRe model constrained by 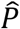 has the minimum AIC among all constrained RelRe models, and no other constrained RelRe model has the same AIC as 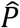. If the beam width, *w*, is set to ∞, then the algorithm outputs a pair of 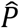 and its AIC as (*P*_*min*_, *AIC*_*min*_) when FindPart-*w* terminates.

**Proof**. From Lemma 4, *S*_*h*_ contains a refinement of 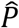 for each 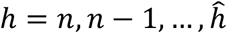. Furthermore, all partitions in *S*_*h*_ are rank-*h* partitions. Since 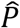 itself is the refinement of 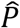 and 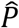 is a rank-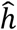 partition, 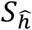 contains 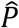. When 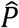 is evaluated as 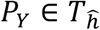 in the rank-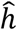 loop, 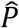 and its AIC is recorded as (*P*_*min*_, *AIC*_*min*_), and *P*_*min*_ and *AIC*_*min*_ will not be updated in the lower rank afterwards, because 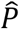 has the only partition that has the smallest AIC among all partitions of *V*. As a result, the algorithm outputs a pair of 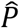 and its AIC as (*P*_*min*_, *AIC*_*min*_) when it terminates. □

Theorem 1 assumes that there is only one constrained RelRe model that has the minimum AIC. If multiple constrained RelRe models have the minimum AIC, then the algorithm outputs the constraining partition and AIC of one of such models. However, such a case seems quite rare.

### Relationship between Algorithm FindPart-1 and Hierarchical Clustering

Clustering is a data analysis technique widely used in various research fields (47). The FindPart-1 algorithm is closely related to the hierarchical clustering algorithm (48), which is one of the most popular clustering algorithms. Hierarchical clustering uses a scoring function to define the goodness of a clustering result, and AIC can be used as the scoring function of hierarchical clustering. When we apply hierarchical clustering to a set of *n* lineages, the hierarchical clustering starts with *n* singleton clusters, each of which consists of a single lineage. These initial *n* singleton clusters at the beginning of hierarchical clustering correspond to the finest partition in the FindPart-1 algorithm. Then, hierarchical clustering iteratively merges the best pair of clusters until the number of clusters becomes one. An iterative step with *h* clusters in hierarchical clustering receives the best clustering result with *h* + 1 clusters determined in the previous step. This iterative step in hierarchical clustering corresponds to a step in the main loop with rank *h* in FindPart-1. Hierarchical clustering evaluates all 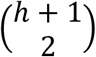 clustering results obtained by merging a pair of *h* + 1 clusters at an iterative step with *h* clusters. When we find the best clustering result with hierarchical clustering, the clustering results with *h* = *n, n* − 1, …,1 were compared using the scoring function. Therefore, the total number of maximum likelihood estimations required to evaluate clustering results with their AIC values is given by

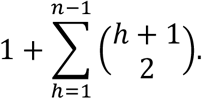

Similarly, FindPart-1 generates 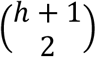 immediate aggregations of the rank-(*h* + 1) partition at an iterative step with a rank-*h* partition. However, FindPart-1 evaluates only a subset of these immediate aggregations by skipping any partition that is an aggregation of a partition in *U* according to Lemma 3. The total number of maximum likelihood estimations needed with FindPart-1 is smaller than that of hierarchical clustering, and the reduction in the number of maximum likelihood estimations is purely attributed to the use of Lemma 3.

### Parameter Estimation and Model Selection Using Simulated Data

#### Hypothetical Observation Dataset of Lineage Replacement

To evaluate the performance of the FindPart-*w* with different beam width *w*, we created hypothetical observation datasets of lineage replacement using simulations. In each simulation, we set the start date of observation, *t*_*s*_, to an arbitrary date, and the end date of observation, *t*_*e*_, to 90 days after *t*_*s*_, so that the length of the observation period was almost the same as that of the actual dataset, which will be described in another section. Assuming all lineages existed at the start of observation, we set all 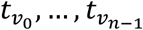 to *t*_*s*_.

For a given number of lineages *n*, we generate true parameters (*ρ*_0_, …, *ρ*_*n*−1_) and 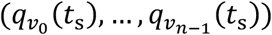 of the true model as follows. For simplicity, rank *h*, the number of unique values in (*ρ*_0_, …, *ρ*_*n*−1_), in the true model was set to the closest integer to (2*n*/3) for every simulation. The value of *ρ*_0_ was always set to 1.0. To generate values of (*ρ*_1_, …, *ρ*_*n*−1_), we first created a temporary *h*-length vector 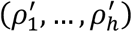, where the first element 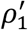 was fixed to 1.0 and the remaining *h* − 1 elements 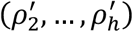 were randomly sampled from a uniform distribution from 0.9 to 2.0. This range of uniform distribution was selected according to the relative effective reproduction numbers found by previous studies of SARS-CoV-2 lineages (18, 23, 25, 32). The values of (*ρ*_1_, …, *ρ*_*n*−1_) were randomly sampled with replacement from the temporary vector 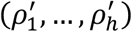 until (*ρ*_0_, …, *ρ*_*n*−1_) contained exactly *h* unique values. Note that the above procedure gives a subject lineage *v*_*i*_ (*i* ≠ 0) a chance to have the same *R*_*RI*_ as the baseline *v*_0_. From the generated parameters, (*ρ*_0_, …, *ρ*_*n*−1_), the true partition was determined so that any block contains two lineages *v*_*i*_, *v*_*j*_ if and only if *ρ*_*i*_ = *ρ*_*j*_.

We generated initial frequencies of the subject lineages 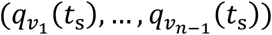 by random sampling from a uniform distribution from 0.01 to 0.1. The value of 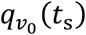) was randomly sampled from a uniform distribution from 0.70 to 0.95. This ensured that the baseline lineage had a higher frequency than the subject lineages. Then, each frequency was divided by the sum of all frequencies to normalize them so that the sum of 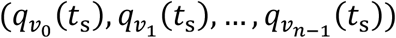 equaled 1.0.

Using parameters 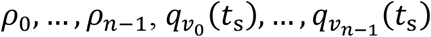 generated above, we conducted a simulation of lineage replacement. For each day *t* from *t*_*s*_ + 1 to *t*_*e*_, we calculated daily frequencies of lineages 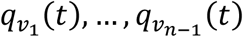 with Equations (9) and 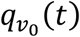 with Equation (10). The approximation, which is described in the subsection of Calculation of the Trajectory of Lineages Replacement, was used to calculate relative frequencies for the first *l* − 1 days. For each day *t* from *t*_*s*_ to *t*_*e*_, observation counts of lineages, 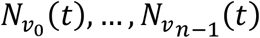, were sampled from a multinomial distribution of size 100 × n of which probabilities were 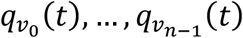.

For each *n* ∈ {5, 10, 15, 20, 25, 30, 35}, we created ten observation datasets by randomly generating parameters of *n* lineages. As a result, a total of 70 observation datasets were obtained and used to evaluate the running time and accuracy of the FindPart-*w* algorithm.

### Parameter Estimation and Model Selection Using FindPart-*w* Algorithm

To evaluate the running time and accuracy of the FindPart-*w* algorithm, we applied the FindPart-*w* algorithm with different *w* for hypothetical observations generated by simulation. For each of the 70 observation datasets, the algorithm was run 6 times by changing *w* to 50, 40, 30, 20, 10, and 1. Due to the long calculation time, FindPart-∞ was able to run with only 40 observation datasets having 5, 10, 15, and 20 lineages, as will be described in the Results section. For each dataset, we recorded the number of performed maximum likelihood estimations and the output (*P*_*min*_, *AIC*_*min*_) of each algorithm. The AIC of the original RelRe models were also calculated and recorded for each dataset for comparison. We did not run the naïve algorithm, and the number of calculations needed for the naïve algorithm was obtained theoretically using the Bell number.

### Model Selection Using *k*-means Algorithm

To evaluate the ability of the FindPart-*w* algorithm at identifying lineages that have the same *R*_*RI*_, we compared the partition found by our algorithm with the clustering result obtained by the *k*-means algorithm (49), which is one of the most well-known clustering algorithms. For each of the 70 observation datasets, the *k*-means algorithm was applied as follows. Let *V* = {*v*_0_, …, *v*_*n*−1_} be a set of lineages. Suppose *ρ*_0_ = 1.0 and *ρ*_1_, …, *ρ*_*n*−1_ are the maximum likelihood estimates of *R*_*RI*_ of *v*_1_, …, *v*_*n*−1_ w.r.t. *v*_0_ estimated under the original RelRe model from the observation dataset, respectively. For each *k* = 1, …, *n*, one-dimensional data *ρ*_0_, …, *ρ*_*n*−1_ were clustered into *k* clusters using *k*-means algorithm. A partition *P*_F_ representing the clustering result was determined so that *v*_*i*_ and *v*_*j*_ belong to the same block in *P*_*C*_ if and only if *ρ*_*i*_ and *ρ*_*j*_ belong to the same cluster. Then, the maximum likelihood estimation of parameters was performed under the RelRe model constrained by *P*_*C*_, and the AIC was determined from the maximum likelihood and the number of free parameters. The *k*-means clustering was performed *n* times using randomly generated initial points, since the results of the *k*-means algorithm depend on initial points. Finally, out of *n*^2^ partitions converted from the clustering results, the partition achieving the minimum AIC was determined and recorded with its AIC. The *k*-means clustering was performed using the Clustering package (version 0.15.7) in the Julia language (version 1.11.1).

### Evaluation of the Ability to Identify Lineages Sharing the Same *R*_*t*2_

For each hypothetical observation dataset, we compared the partition output by the algorithm with the true partition converted from the parameters that were used to generate the hypothetical observation dataset. The comparison was done in a pairwise manner as described below.

Let *V* = {*v*_0_, …, *v*_*n*−1_} be a set of *n* lineages. For each combination of two lineages (*v*_*i*_, *v*_*j*_), where *i* < *j*, we checked whether *v*_*i*_ and *v*_*j*_ belonged to the same block in the true partition and the predicted partition. If *v*_*i*_ and *v*_*j*_ belonged to the same block in the true partition and the predicted partition, then this pair was considered as a true positive (TP). If *v*_*i*_ and *v*_*j*_ belonged to the same block in the true partition and belonged to different blocks in the predicted partition, then this pair was considered as a false negative (FN). If *v*_*i*_ and *v*_*j*_ belonged to different blocks in the true partition and belonged to different blocks in the predicted partition, then this pair was considered as a true negative (TN). If *v*_*i*_ and *v*_*j*_ belonged to different blocks in the true partition and belonged to the same block in the predicted partition, then this pair was considered as a false positive (FP). Thus, for a given true partition and a given predicted partition, a total of *n* (*n* − 1)/2 pairs of lineages were compared, and the summation of TP, FN, TN, and FP equaled to this number.

We evaluated the ability of each algorithm at finding lineages that shared identical *R*_*RI*_ using sensitivity, specificity, and precision (positive predictive value) (50). These metrics were calculated using the following equations:

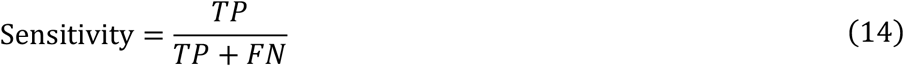

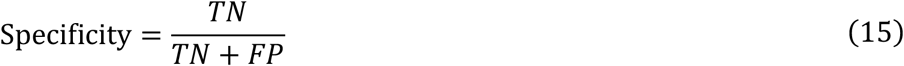

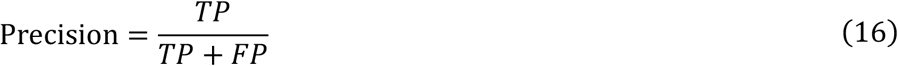

### Parameter Estimation and Model Selection Using Actual Dataset

#### Data Collection

We downloaded nucleotide sequences and their metadata of human SARS-CoV-2 collected in the United States of America (USA) from the GISAID database (51). We used metadata of nucleotide sequences collected during the period from 1^st^ October 2024 to 31^st^ December 2024 and submitted to GISAID no later than 31^st^ January 2025. We set the start date of our analysis, *t*_*s*_, to 1^st^ October 2024 and the end date, *t*_*e*_, to 31^st^ December 2024. For each date from *t*_*s*_ to *t*_*e*_, we counted the Pango lineage names of sequences collected on that date. We excluded sequences of which Pango lineage names were unassigned. To identify major lineages during the analysis period, we first normalized the daily counts of lineages into daily relative frequencies using the daily count of all lineages. Then, the lineages were sorted in descending order based on the sum of relative frequencies during the analysis period. According to this order, the lineages were selected as the major lineages until the total sequence count of the selected lineage reached 90 percent. As a result, we obtained a dataset that contained 18,556 sequences consisting of 19 major lineages. The daily count data of major lineages were then used for all subsequent downstream analysis.

#### Parameter Estimation and Model Selection

To find SARS-CoV-2 lineages that share the same *R*_*RI*_, we applied the FindPart-*w* algorithm to the actual observation data. We used KP.3.1.1 as the baseline, as it was the most abundant lineage at the start date of our analysis.

First, we conducted a maximum likelihood estimation under the original RelRe model using the observation dataset. We recorded the AIC of the original RelRe model and the maximum likelihood estimates of parameters 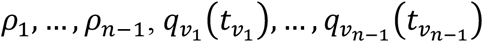. Second, we applied the FindPart-50 algorithm to find the partition achieving the smallest AIC. The partition and AIC output by the FindPart-50 algorithm were recorded, and the AIC was compared with that of the original RelRe model.

#### Implementation and Computing Environment

The algorithm FindPart-*w* (Figure 2) was implemented using the Julia language (version 1.11.1). The maximum likelihood estimation under constrained RelRe models was also implemented using the Julia language, by modifying the original RelRe program (52). In the modified version of RelRe program, we introduced an optional command-line argument that allowed us to give an RGS to represent a partition of lineages sharing the same reproduction numbers. The program codes of the constrained version of the RelRe program and the FindPart-*w* are available at another GitHub repository (53). We used R version 4.4.2 for statistical tests and the visualisation of results. Computational experiments using hypothetical dataset were performed on the Grand Chariot computing nodes in the supercomputer system at Hokkaido University (54). Each experiment using hypothetical observation data was performed used a computing node equipped with a total memory of 384 GB, with two sockets of Intel (R) Xeon (R) Processor Gold 6148 processor where each socket has 20 cores running at a maximum clock speed of 3.7 GHz. Analysis using actual observation data was performed using a MacBook Pro with a total memory of 64 GB, with 2.4 GHz 8-Core Intel Core i9 processor running at a maximum clock speed of 2667 MHz.

## Results

### Performance on the Simulated Data

#### Running Time

Theorem 1 ensures that the FindPart-*w* algorithm finds the partition that achieves the smallest AIC among all partitions when there is no restriction on the beam width (*w* = ∞). We first evaluated the effect of *w* on the running time of the algorithm using simulated datasets.

For a given dataset, total running time is correlated with the number of maximum likelihood estimations (Figure S1). This means that the running time of the maximum likelihood estimation does not largely differ from one partition to another, and the total running time of the algorithm depends mostly on the number of performed maximum likelihood estimations. For this reason, we evaluate the running time of the algorithm using the number of performed maximum likelihood estimations.

Figure 3 shows the relationship between the number of lineages and the number of maximum likelihood estimations performed by the Naïve search, FindPart-∞, FindPart-50, FindPart-40, FindPart-30, FindPart-20, FindPart-10, and FindPart-1 when they were applied to the simulated datasets. Note that the number of maximum likelihood estimations on the y-axis is log-10 transformed. A straight line on this semi-log plot indicates exponential growth, a convex line indicates super-exponential growth, and a concave line indicates sub-exponential growth.

**Figure 3.**
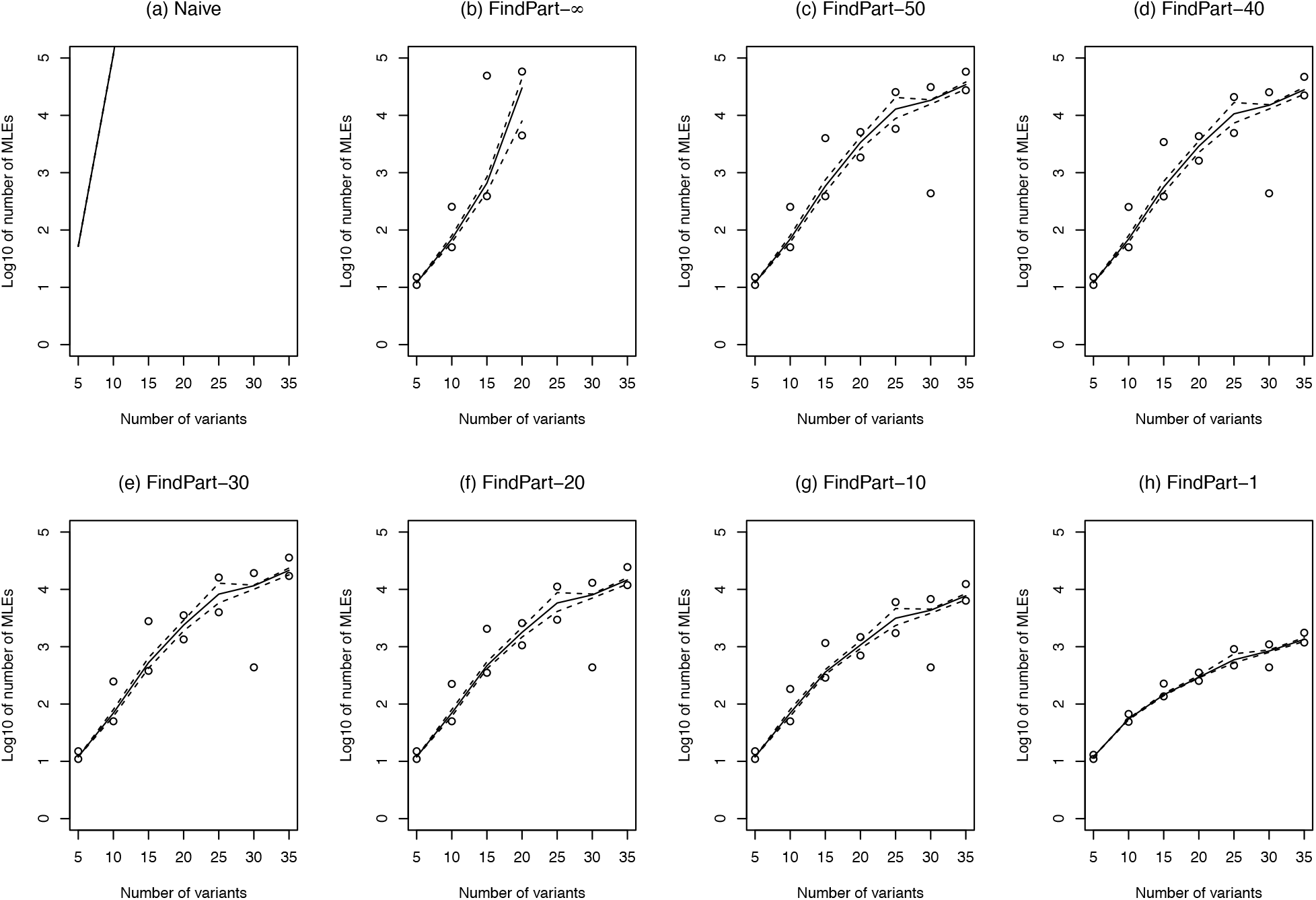
Relationship between the number of lineages and the number of maximum likelihood estimations required by (a) Naïve, performed by (b) FindPart-∞, (c) FindPart −50, (d) FindPart-40, (e) FindPart-30, (f) FindPart-20, (g) FindPart-10, and (h) FindPart-1. In each panel, the x-axis represents the number of lineages, and the y-axis represents the log-10 of the number of maximum likelihood estimations (MLEs). The solid lines represent the median, the dashed lines represent the first and third quantiles, and the circles represent the minimum and maximum across ten simulated datasets. The number of calculations required by the Naïve algorithm was determined by the Bell number, and quantiles and maximum and minimum were not shown in (a).

The FindPart-∞ is guaranteed to find the constraining partition of the best constrained RelRe model having minimum AIC for any given set of lineages (Theorem 1). This is also true for the Naïve search, which calculates AICs for all possible constrained RelRe models. Using Lemma 3, FindPart-*w* skips the maximum likelihood estimation of RelRe models that have no chance of having minimum AIC. If FindPart-∞ does not skip maximum likelihood estimations of any constrained RelRe model using Lemma 3, FindPart-∞ checks all possible constrained RelRe models. In this case, the number of maximum likelihood estimations of FindPart-∞ is equal to that of the Naïve search. Comparing the numbers of maximum likelihood estimations of FindPart-∞ to those of Naïve search, we can quantify the effect of using Lemma 3 on the number of maximum likelihood estimations. From Figure 3 (a) and (b) and Table 1, one can find that FindPart-∞ has a significant improvement over the Naïve search. However, FindPart-∞ was unable to finish experiments when lineage sizes are larger than 20, due to the long computational times resulting from super-exponential growth in the number of maximum likelihood estimations (Figure 3 (b)).

**Table 1.** Effect of Lemma 3 on the Number of Maximum Likelihood Estimations.

| Number of lineages (n) | FindPart-∞ | | | Naive | p-value |
| --- | --- | --- | --- | --- | --- |
|  | Minimum | Mean | Maximum |  |  |
| 5 | 11 | 12.2 | 15 | 52 | $8.57 \times 10^{-16}$ |
| 10 | 50 | 99.6 | 254 | $1.16 \times 10^5$ | $2.93 \times 10^{-30}$ |
| 15 | 389 | 5695.8 | 49198 | $1.38 \times 10^9$ | $4.01 \times 10^{-46}$ |
| 20 | 4457 | 28734.4 | 58044 | $5.17 \times 10^{13}$ | $4.29 \times 10^{-8}$ |
| 25 | n.a. | n.a. | n.a. | $4.64 \times 10^{18}$ | n.a. |
| 30 | n.a. | n.a. | n.a. | $8.47 \times 10^{23}$ | n.a. |
| 35 | n.a. | n.a. | n.a. | $2.87 \times 10^{29}$ | n.a. |

The minimum, mean, and maximum values for FindPart- ∞ are based on 10 simulation replicates for each lineage size $n$ . A cell with n.a. indicates missing values where we were unable to finish the calculation. The p-values were calculated with t-tests with the null hypothesis that the mean of the number of maximum likelihood estimations in FindPart- ∞ is the number of maximum likelihood estimations under Naïve search.

To reduce computational time, we restricted the beam width, *w*, to be 50, 40, 30, 20, 10, and 1. All of FindPart-50, FindPart-40, FindPart-30, FindPart-20, FindPart-10, and FindPart-1 showed concave lines in their log-linear plots, indicating the sub-exponential growth in the number of maximum likelihood calculations (Figure 3 (c) to (h)). The restriction on the beam width contributed to reducing the number of maximum likelihood estimations significantly. Raw values are given in Table S1.

Table 2 compares the number of maximum likelihood estimations needed by the FindPart-1 with that by hierarchical clustering. As discussed in a subsection of Materials and Methods, the reduction in the number of maximum likelihood estimations from hierarchical clustering to that of the FindPart-1 is purely attributed to the use of Lemma 3. From the p-values of the t-test shown in Table 2, we can conclude that the number of maximum likelihood estimations in FindPart-1 these significantly smaller than that in hierarchical clustering. Raw values are given in Table S1.

**Table 2.**
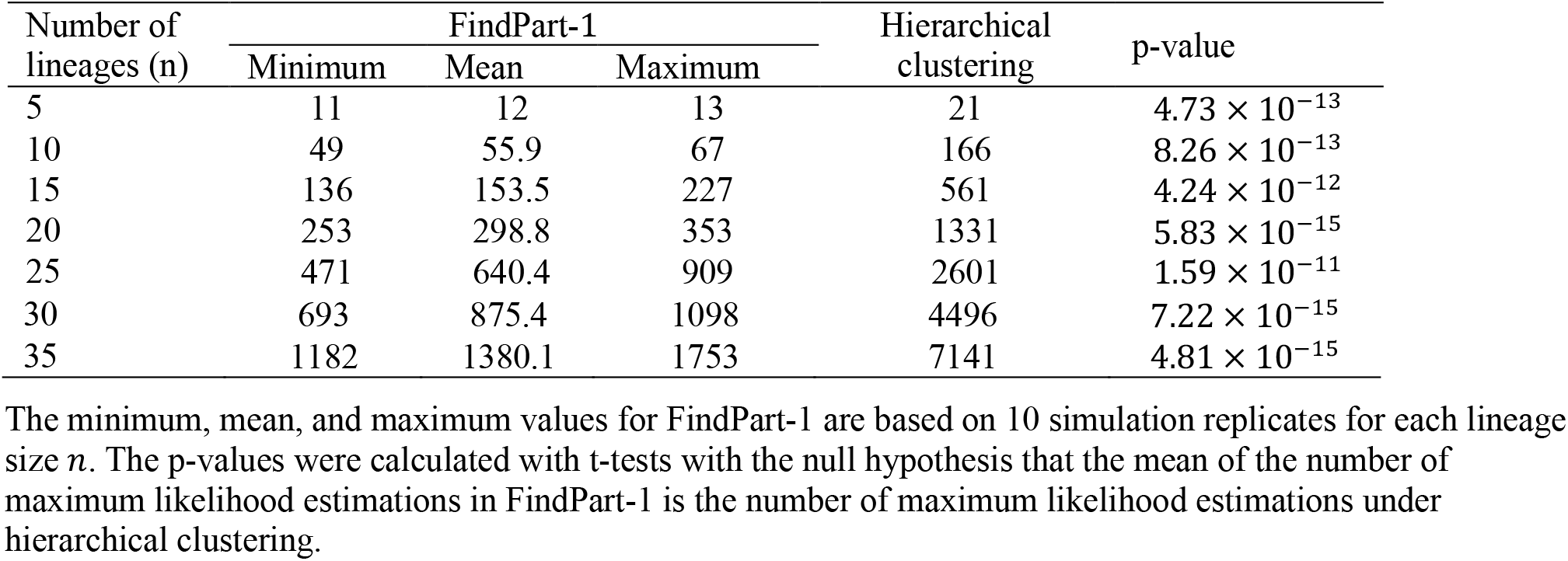
Effect of Lemma 3 on the Number of Maximum Likelihood Estimations.

### Effect of the Beam Width on the AIC of Output Constrained RelRe Models

Next, we investigated the effect of the beam width on the AIC of RelRe models constrained by the partition output by the FindPart-*w* algorithm. For each observation dataset created by simulation, we recorded the AIC values of RelRe models constrained by the partitions output by FindPart-50, FindPart-40, FindPart-30, FindPart-20, FindPart-10, FindPart-1, *k*-means, and the original RelRe model. Δ-AIC was calculated by subtracting the minimum AIC from the AIC of the RelRe models output by each method (Table 3). A Δ-AIC value of zero indicates that the method found the partition that achieved the minimum AIC among all tested methods. A positive Δ-AIC value indicates how much the AIC of the partition output by a specified method is larger than the minimum AIC. Raw AIC values are provided in Table S2.

**Table 3.** Statistics on the Δ-AIC of partitions output by FindPart-*w* compared to partitions output by FindPart-50.

| Procedure to find the best partition | Statistic | Number of lineages ($n$) | | | | | | |
| --- | --- | --- | --- | --- | --- | --- | --- | --- |
|  |  | 5 | 10 | 15 | 20 | 25 | 30 | 35 |
| FindPart-50 | Minimum | 0.000 | 0.000 | 0.000 | 0.000 | 0.000 | 0.000 | 0.000 |
|  | Mean | 0.000 | 0.000 | 0.000 | 0.000 | 0.000 | 0.000 | 0.000 |
|  | Maximum | 0.000 | 0.000 | 0.000 | 0.000 | 0.000 | 0.000 | 0.000 |
| FindPart-40 | Minimum | 0.000 | 0.000 | 0.000 | 0.000 | 0.000 | 0.000 | 0.000 |
|  | Mean | 0.000 | 0.000 | 0.000 | 0.000 | 0.000 | 0.000 | 0.000 |
|  | Maximum | 0.000 | 0.000 | 0.000 | 0.000 | 0.000 | 0.000 | 0.000 |
| FindPart-30 | Minimum | 0.000 | 0.000 | 0.000 | 0.000 | 0.000 | 0.000 | 0.000 |
|  | Mean | 0.000 | 0.000 | 0.000 | 0.000 | 0.000 | 0.000 | 0.052 |
|  | Maximum | 0.000 | 0.000 | 0.000 | 0.000 | 0.000 | 0.000 | 0.520 |
| FindPart-20 | Minimum | 0.000 | 0.000 | 0.000 | 0.000 | 0.000 | 0.000 | 0.000 |
|  | Mean | 0.000 | 0.000 | 0.000 | 0.000 | 0.011 | 0.000 | 0.056 |
|  | Maximum | 0.000 | 0.000 | 0.000 | 0.000 | 0.089 | 0.000 | 0.520 |
| FindPart-10 | Minimum | 0.000 | 0.000 | 0.000 | 0.000 | 0.000 | 0.000 | 0.000 |
|  | Mean | 0.000 | 0.000 | 0.000 | 0.000 | 0.089 | 0.136 | 0.288 |
|  | Maximum | 0.000 | 0.000 | 0.000 | 0.000 | 0.893 | 1.355 | 0.955 |
| FindPart-1 | Minimum | 0.000 | 0.000 | 0.000 | 0.000 | 0.000 | 0.000 | 0.000 |
|  | Mean | 0.000 | 0.000 | 0.147 | 0.000 | 0.687 | 1.171 | 0.829 |
|  | Maximum | 0.000 | 0.000 | 1.471 | 0.000 | 2.484 | 9.828 | 1.816 |
| (RelRe) | Minimum | 1.516 | 3.890 | 2.872 | 11.170 | 12.084 | 25.797 | 25.928 |
|  | Mean | 3.298 | 5.434 | 8.706 | 16.805 | 23.498 | 30.555 | 45.516 |
|  | Maximum | 4.201 | 7.603 | 12.657 | 22.468 | 31.654 | 41.397 | 66.146 |
| ($k$-means) | Minimum | 0.000 | 0.000 | 0.000 | 0.000 | 0.000 | 0.000 | 0.000 |
|  | Mean | 0.000 | 0.096 | 0.475 | 0.550 | 1.958 | 2.370 | 3.180 |
|  | Maximum | 0.000 | 0.960 | 3.218 | 5.505 | 6.312 | 10.274 | 8.015 |

The minimum, mean, and maximum values for Δ-AIC are based on 10 simulation replicates for each lineage size $n$ .

Since FindPart-∞ is theoretically guaranteed to find the partition that achieves the minimum AIC, it was natural to expect that FindPart-*w* with a larger *w* would output a constraining partition with a smaller Δ-AIC. Consistent with this expectation, the mean Δ-AIC decreased or remained the same when *w* increased for all lineage sizes we tested (Table 3). The maximum of Δ-AIC for FindPart-50 and FindPart-40 was zero for all lineage sizes we tested. This means that partitions found by FindPart-50 and FindPart-40 achieved the minimum AIC among all the tested methods for each observation dataset. When *n* = 5, Δ-AIC was zero for all *w* for all datasets. When *n* = 35, in contrast, the mean Δ-AIC values improved when *w* was increased from 1 to 10, from 10 to 20, and from 30 to 40, and no improvement on Δ-AIC was observed when *w* was increased from 40 to 50. These results suggested that the beam width *w* required to get a converged value of AIC increased as the number of lineages *n* increased. Importantly, for all *n* we tested, the mean Δ-AIC of the RelRe models constrained by partitions found by FindPart-1 was smaller than 2. This suggested that the partitions found by FindPart-1 were as good as the partitions found by FindPart-50 (and FindPart-40) in terms of AIC (55).

Δ-AIC values of the original RelRe models in Table 3 represent the amount of AIC decreased in the RelRe model constrained by the partition found by FindPart-50 (and FindPart-40) compared to the original RelRe models. When *n* ≥ 10, the mean Δ-AIC of the original RelRe models was greater than 4. This suggested that the original RelRe models had considerably less support than the RelRe models constrained by partitions found by FindPart-50 (and FindPart-40) in terms of AIC (55).

To compare the performance of FindPart-*w* algorithm to an existing clustering algorithm, the clustering result of the *k*-means algorithm was included in Table 3. For all lineage sizes *n* ≥ 10 we tested, the clustering results of the *k*-means were not as good as the partitions found by the FindPart-*w* algorithms in terms of AIC. When *n* ≥ 30, the mean Δ-AIC of the RelRe models constrained by the clustering result of the *k*-means was greater than 2.0. This suggested that when *n* ≥ 30 the clustering result of the *k*-means had substantially less support than partitions found by FindPart-50 (and FindPart-40) in terms of AIC (55).

### Ability to Find the Lineages Sharing the Same *R*_*t*2_

To evaluate the ability of the FindPart-*w* algorithm to find lineages with the same *R*_*RI*_, we compared the partition output by the algorithm with the true partition determined from *ρ*_0_, …, *ρ*_*n*−1_, with which observation datasets were generated.

Table 4 shows the mean sensitivity, mean specificity, and mean precision of FindPart-50, FindPart-20, and FindPart-1 in finding a pair of lineages having the same *R*_*RI*_ across ten hypothetical datasets for each lineage size *n*. Raw values are provided in Table S3.

**Table 4.** Sensitivity, specificity, and precision of FindPart-*w* at identifying lineages that share the same *R*_*RI*_.

| Metric | Method | Number of lineages (n) |  |  |  |  |  |  |
| --- | --- | --- | --- | --- | --- | --- | --- | --- |
|  |  | 5 | 10 | 15 | 20 | 25 | 30 | 35 |
| Mean sensitivity | FindPart-50 | <u>0.950</u> | <u>0.825</u> | <u>0.645</u> | <u>0.746</u> | 0.675 | <u>0.786</u> | 0.755 |
|  | FindPart-20 | <u>0.950</u> | <u>0.825</u> | <u>0.645</u> | <u>0.746</u> | <u>0.685</u> | <u>0.786</u> | 0.755 |
|  | FindPart-1 | <u>0.950</u> | <u>0.825</u> | 0.605 | <u>0.746</u> | 0.654 | 0.770 | <u>0.760</u> |
| | ($k$-means) | (0.950) | (0.850) | (0.620) | (0.759) | (0.611) | (0.776) | (0.746) |
| Mean specificity | FindPart-50 | <u>1.000</u> | <u>0.983</u> | <u>0.976</u> | <u>0.977</u> | 0.967 | <u>0.976</u> | <u>0.979</u> |
|  | FindPart-20 | <u>1.000</u> | <u>0.983</u> | <u>0.976</u> | <u>0.977</u> | 0.967 | <u>0.976</u> | 0.977 |
|  | FindPart-1 | <u>1.000</u> | <u>0.983</u> | 0.975 | <u>0.977</u> | <u>0.970</u> | 0.974 | 0.977 |
| | ($k$-means) | (1.000) | (0.983) | (0.976) | (0.976) | (0.975) | (0.979) | (0.980) |
| Mean precision | FindPart-50 | <u>1.000</u> | <u>0.813</u> | <u>0.656</u> | <u>0.633</u> | 0.452 | <u>0.506</u> | <u>0.491</u> |
|  | FindPart-20 | <u>1.000</u> | <u>0.813</u> | <u>0.656</u> | <u>0.633</u> | <u>0.456</u> | <u>0.506</u> | 0.475 |
|  | FindPart-1 | <u>1.000</u> | <u>0.813</u> | 0.631 | <u>0.633</u> | 0.448 | 0.482 | 0.477 |
| | ($k$-means) | (1.000) | (0.828) | (0.642) | (0.611) | (0.508) | (0.543) | (0.503) |

Maximum values among FindPart-50, FindPart-20, and FindPart-1 for each combination of lineage size and metric were underlined.

The mean sensitivity of FindPart-50, FindPart-20, and FindPart-1 ranged from 0.605 to 0.950. Unlike performance measured by the AIC, the mean sensitivity of FindPart-50 was not always better than that of FindPart-20, and FindPart-1. FindPart-50 achieved the best performance in terms of mean sensitivity when *n* = 5, 10, 15, 20, and 30. However, FindPart-20 outperformed FindPart-50 in terms of mean sensitivity when *n* = 25, and FindPart-1 outperformed FindPart-50 in terms of mean sensitivity for *n* = 35. The sensitivities observed across 70 datasets did not differ significantly between FindPart-50 and FindPart-20 (*p* = 1.000 with paired Wilcoxon test), and they also did not differ significantly between FindPart-50 and FindPart-1 (*p* = 0.255 with paired Wilcoxon test).

The mean specificities for FindPart-50, FindPart-20, and FindPart-1 ranged from 0.967 to 1.000 across different sizes of *n*. The mean specificity of FindPart-50 was equal to that of FindPart-20 except when *n* = 35. The mean specificity of FindPart-50 was equal to that of FindPart-1 when *n* = 5, 10, and 20, but FindPart-1 showed slightly lower mean specificity than FindPart-50 when *n* = 15, 30, and 35. The mean specificity of FindPart-1 was slightly higher than that of FindPart-50 and FindPart-20, when *n* = 25. The specificities observed across 70 datasets did not differ significantly between FindPart-50 and FindPart-20 (*p* = 0.371 with paired Wilcoxon test), and they also did not differ significantly between FindPart-50 and FindPart-1 (*p* = 0.414 with paired Wilcoxon test).

The mean precision for FindPart-50, FindPart-20, and FindPart-1 ranged from 0.448 to 1.000. In general, the mean precision decreased as the number of lineages increased. For instance, the mean precision for *n* = 5, was 1.000 for FindPart-50, FindPart-20, and FindPart-1, but when *n* ≥ 25, it was dropped to around 0.500 for all three methods. The precision observed across 70 datasets did not differ significantly between FindPart-50 and FindPart-20 (*p* = 0.789 with paired Wilcoxon test), and they also did not differ significantly between FindPart-50 and FindPart-1 (*p* = 0.085 with paired Wilcoxon test).

To evaluate the ability of FindPart-*w* algorithms to find two lineages having the same *R*_*RI*_, we included results obtained by the *k*-means algorithm in Table 4. The sensitivities observed across 70 datasets did not differ significantly between FindPart-50 and *k*-means (*p* = 0.520 with paired Wilcoxon test). The specificities observed across 70 datasets differed significantly between FindPart-50 and *k*-means (*p* = 0.023 with paired Wilcoxon test). The precision observed across 70 datasets did not differ significantly between FindPart-50 and *k*-means (*p* = 0.298, with paired Wilcoxon test). These results suggested that FindPart-50 had a similar ability to find two lineages that shared the same *R*_*RI*_ as *k*-means, but *k*-means had a better ability to find two lineages that did not share the same *R*_*RI*_ than FindPart-50. The reason for the significant difference in the specificity between FindPart-50 and *k*-means will be discussed in the Discussion section.

### Applications to the Actual Dataset from the United States of America

Using actual sequence count data of SARS-CoV-2 lineages from the United States, we searched for the best constrained RelRe model by the FindPart-*w* algorithm with a beam width *w* = 1. We also conducted the maximum likelihood estimation of parameters under the original RelRe model. The maximum likelihood estimation of parameters under the original RelRe model took about 46 seconds without 95% C.I. calculations and about 151seconds with 95% C.I. calculations using a MacBook Pro (see the Implementation and Computing Environment Subsection in Methods for its specification). FindPart-1 took about 29 minutes with a total of 558 maximum likelihood estimations (without 95% C.I. calculations) to find the partition of the best constrained RelRe model using the same MacBook Pro. The 95% C.I. of parameters under the constrained RelRe models were calculated only for the best constrained RelRe model, and it took about 99 seconds using the same MacBook Pro.

Table 5 compares the AIC of the constrained RelRe model found by the FindPart-1 algorithm with that of the original RelRe model. The actual sequence count data we used contained 19 SARS-CoV-2 lineages (*n*). The original RelRe model had a total of 2*n* − 2 = 36 free parameters with a maximum log-likelihood of –3422.47, and the AIC of the original RelRe model was calculated to be 6916.95. The rank (*h*) of the partition that was output by FindPart-1 was 10. The RelRe model constrained by this partition had a total of *n* + *h* − 2 = 27 free parameters with a maximum likelihood of –3423.94, and the AIC of the constrained RelRe model was calculated to be 6901.88. The AIC of the constrained RelRe model found by FindPart-1 was 15.07 smaller than the original model. This result showed that the constrained RelRe model found by FindPart-1 was significantly better than the original RelRe model in terms of AIC, because Δ-AIC > 10 (55).

**Table 5.** Comparison of the original RelRe model and the constrained RelRe model found by the FindPart-1 algorithm.

| Model name | The number of lineages ($n$) | Maximum log-likelihood ($\ln(\hat{L})$) | Rank of the partition of lineages ($h$) | The number of free parameters ($k$) | AIC | $\Delta\text{-AIC}$ |
| --- | --- | --- | --- | --- | --- | --- |
| Original RelRe model | 19 | $-3422.47$ | 19 | 36 | 6916.95 | 15.07 |
| Constrained RelRe model found by FindPart-1 | 19 | $-3423.94$ | 10 | 27 | 6901.88 | 0.000 |

Table 6 shows a list of 19 lineages in the actual dataset along with their maximum likelihood estimates of *R*_*RI*_ w.r.t. KP.3.1.1, which were estimated under the original RelRe model. LP.8.1 had the highest estimates of *R*_*RI*_ at 1.239, and KP.2.3 had the lowest estimates of *R*_*RI*_ at 0.899. Some lineages had very similar values of *R*_*RI*_. For example, the lineages MC.1, JN.1.8, JN.1.40 had *R*_*RI*_ of 1.056, 1.050, 1.047, respectively. There was an overlap among the 95 % confidence intervals (C.I.) of the *R*_*RI*_ of these three lineages. Similarly, the lineages JN.1.11, KP.1.1.3, JN.1.16.1, JN.1 had *R*_*RI*_ of 1.035, 1.032, 1.030, 1.025, and 1.025, respectively, and there was an overlap among the 95 % C.I. of the *R*_*RI*_ of these five lineages. These results suggested that there was a possibility that some lineages shared the same *R*_*RI*_.

**Table 6.** A list of 19 lineages in the actual dataset and maximum likelihood estimates of their *R*_*RI*_ w.r.t. KP.3.1.1 under the original RelRe model.

| lineages | $R_{RI}(\rho)$ | 95% C.I. of $R_{RI}(\rho)$ |
| --- | --- | --- |
| LP.8.1 | 1.239 | 1.233–1.245 |
| XEC | 1.092 | 1.089–1.094 |
| JN.1.18.6 | 1.076 | 1.071–1.079 |
| MC.1 | 1.056 | 1.051–1.062 |
| JN.1.8 | 1.050 | 1.047–1.057 |
| JN.1.40 | 1.047 | 1.040–1.052 |
| JN.1.11 | 1.035 | 1.027–1.037 |
| KP.1.1.3 | 1.032 | 1.026–1.036 |
| JN.1.16 | 1.030 | 1.025–1.034 |
| JN.1.16.1 | 1.025 | 1.020–1.032 |
| JN.1 | 1.025 | 1.008–1.028 |
| MB.1.1 | 1.015 | 1.000–1.025 |
| KP.3.1 | 1.008 | 1.004–1.011 |
| KP.3.1.1 | 1.000 | 1.000–1.000 |
| KP.3 | 0.990 | 0.983–0.996 |
| XDY | 0.986 | 0.979–0.998 |
| KP.3.3 | 0.983 | 0.970–1.000 |
| KP.2 | 0.949 | 0.938–0.961 |
| KP.2.3 | 0.899 | 0.888–0.907 |

The 19 major lineages in the actual observation dataset from the United States were grouped into 10 blocks by FindPart-1. Table 7 shows the lineages contained in each block, along with the maximum likelihood estimate of their *R*_*RI*_ w.r.t. KP.3.1.1 under the constrained RelRe model found by FindPart-1. Lineages JN.1.40, JN.1.8, and MC.1, which had similar estimates with overlapping 95% C.I. under the original RelRe model (Table 6), were found to share the same *R*_*RI*_ of 1.052 under the constrained RelRe model found by FindPart-1. Similarly, JN.1, JN.1.11, JN.1.16, JN.1.16.1, and KP.1.1.3, which had similar estimates of *R*_*RI*_ with overlapping 95% C.I. under the original RelRe model, were found to share the same *R*_*RI*_ of 1.030 under the constrained RelRe model found by FindPart-1. In contrast to the results obtained under the original RelRe model (Table 6), there was no overlap in the 95% C.I. of *R*_*RI*_ estimated under the constrained RelRe model found by FindPart-1 for any combination of two blocks (Table 7).

**Table 7.** The partition of 19 lineages output by the FindPart-1 and maximum likelihood estimates of their *R*_*RI*_ w.r.t. KP.3.1.1 under the RelRe model constrained by the partition.

| Blocks | $R_{RI}(\rho)$ | 95% C.I. of $R_{RI}$ |
| --- | --- | --- |
| LP.8.1 | 1.236 | 1.223–1.242 |
| XEC | 1.092 | 1.091–1.094 |
| JN.1.18.6 | 1.075 | 1.071–1.080 |
| JN.1.40, JN.1.8, MC.1 | 1.052 | 1.051–1.054 |
| JN.1, JN.1.11, JN.1.16, JN.1.16.1, KP.1.1.3 | 1.030 | 1.028–1.033 |
| KP.3.1, MB.1.1 | 1.008 | 1.006–1.011 |
| KP.3.1.1 | 1.000 | 1.000–1.000 |
| KP.3, KP.3.3, XDY | 0.987 | 0.985–0.993 |
| KP.2 | 0.949 | 0.938–0.961 |
| KP.2.3 | 0.899 | 0.890–0.906 |

Since full Pango lineage names reflect the genetic relationship among lineages, Table 7 allows us to know whether or not lineages with similar genetic background have the same *R*_*RI*_. In some cases, lineages with similar genetic backgrounds shared the same transmissibility. For instance, JN.1.11 (alias of B.1.1.529.2.86.1.1.11) and JN.1.16 (alias of B.1.1.529.2.86.1.1.16), both derived from JN.1 (alias of B.1.1.529.2.86.1.1) were found to share the same *R*_*RI*_ as JN.1, suggesting that genetic differences among these lineages did not alter the transmissibility of the virus. In some cases, lineages with similar genetic backgrounds were found to have different transmissibility. For example, lineages KP.3.1 (alias of B.1.1.529.2.86.1.1.11.1.3.1) and KP.3.1.1 (alias of B.1.1.529.2.86.1.1.11.1.3.1.1), both derived from KP.3 (alias of B.1.1.529.2.86.1.1.11.1.3) are genetically similar, yet they had different *R*_*RI*_, suggesting that closely related lineages can have differences in their transmissibility. In some other cases, lineages with less similar genetic backgrounds were found to share the same *R*_*RI*_. For instance, KP.3.1 (alias of B.1.1.529.2.86.1.1.11.1.3.1) and MB.1.1 (alias of B.1.1.529.2.86.1.1.49.1.1.1) diverged from JN.1 (alias of B.1.1.529.86.1.1). After the divergence, KP.3.1 and MB.1.1 evolved independently, such that both were assigned new lineage names four times in the Pango lineage system. Despite relatively large genetic differences, KP.3.1 and MB.1.1 were found to have the same transmissibility.

## Discussion

This study focused on the problem of identifying groups of lineages that share the same *R*_*RI*_. We introduced a new lineage replacement model, called the constrained RelRe model, by extending the original RelRe model so that some lineages are constrained to share the same *R*_*RI*_ as other lineages. For a given observation dataset, groups of lineages that share the same *R*_*RI*_ can be identified by selecting the constrained RelRe model of which AIC is the minimum among all constrained RelRe models. Equality constraints in a constrained RelRe model can be uniquely represented with a partition of *n* lineages where lineages in the same block share the same *R*_*RI*_. Since the number of possible partitions of *n* lineages is equal to the (*n* − 1)th Bell number, it is computationally impractical to find the partition achieving the minimum AIC by computing the AIC for all possible partitions. To solve this problem, we developed the FindPart-*w* algorithm, which employed two mechanisms to reduce the computational time. The first mechanism takes advantage of Lemma 3, which allows us to determine that a partition has no possibility of achieving the minimum AIC without calculating its maximum likelihood. The second mechanism takes a greedy approach by restricting the beam width *w*, which is the number of partitions passed from one iteration step to the next step in the main loop of the algorithm. Using hypothetical observation datasets created by simulations, we showed that the number of maximum likelihood calculations needed by FindPart-*w* grew sub-exponentially against the number of lineages when *w* < 50, and that FindPart-*w* could find a significantly better model compared to the original RelRe model. When applied to an actual observation dataset of SARS-CoV-2 lineages from the United States, FindPart-1 found a constrained RelRe model that was significantly better than the original RelRe model in terms of AIC. The constrained RelRe model found by FindPart-1 grouped 19 lineages into 10 blocks of lineages having the same *R*_*RI*_, suggesting that several lineages found in the actual dataset shared the same *R*_*RI*_, although they were assigned to different lineage names by the Pango lineage system.

Using actual observation data of lineage counts in the United States, we found that there was a constrained RelRe model of which AIC was significantly smaller than that of the original RelRe model (Table 5). Furthermore, some of the lineages that were identified to share the same transmissibility were found to be genetically closely related according to their Pango lineage names (Table 7). These observations mean that the Pango lineage nomenclature system may have given different lineage names to SARS-CoV-2 strains even if they have the same transmissibility and similar genetic backgrounds. This highlights the possibility of developing a nomenclature system that designates lineages by considering both genetic information and viral transmissibility. This study contributes to the future development of such a nomenclature system by showing that viruses sharing the same transmissibility can be identified from count data of viral sequences. However, the current form of FindPart-*w* relies on the Pango lineage system to get temporal data on the lineage counts. Therefore, future studies should consider developing a method of lineage designation system that does not rely on the Pango lineage system.

Table 1 shows that FindPart-∞ required fewer maximum likelihood calculations than Naïve search for lineage size *n* = 5, 10, 15, 20. Table 2 shows that FindPart-1 required fewer maximum likelihood calculations than hierarchical clustering for lineage size *n* = 5,10,15,20,25,30,35. These results suggested that Lemma 3 has a significant effect on reducing the number of maximum likelihood calculations for the FindPart-*w* algorithm when *w* = ∞ or 1. We did not test effect of Lemma 3 on the number of maximum likelihood estimations of FindPart-*w* for other *w* than ∞ or 1. This is because the number of maximum likelihood estimations without use of Lemma 3 can be calculated analytically only for *w* = ∞ or 1, while knowing the numbers for other *w* than ∞ or 1 without use of Lemma 3 required long computational time. However, we expect that Lemma 3 reduced the number of maximum likelihood estimations in the same way as Table 1 and Table 2 when *w* is other than ∞ or 1.

We can think of FindPart-*w* as a clustering algorithm. FindPart-*w* can be applicable to other kinds of datasets when one performs the clustering analysis using AIC as the scoring function. Theorem 1 ensures that the FindPart-*w* algorithm finds the constraining partition of the RelRe model that achieves the smallest AIC among all partitions when there is no restriction on the beam width (*w* = ∞). This theorem can be generalized to deal with other datasets in a straightforward way, because Lemma 3 uses general properties on the refinement-aggregation relationship of partitions and their AIC. Although Lemma 3 reduced the number of maximum likelihood estimations significantly compared to the Naïve algorithm, further reduction in the number of maximum likelihood calculations was required for practical application. For this reason, we restricted the beam width *w* of FindPart-*w*, which is the number of partitions passed from one iteration step to the next step in the main loop of the algorithm. As described in the Method section, FindPart-1 works almost in the same way as the hierarchical clustering algorithm (48), except calculations of AIC for some partitions can be skipped using Lemma 3. Therefore, one can speed up hierarchical clustering by using FindPart-1 when one performs the clustering analysis using AIC as the scoring function.

In this study, we used the constrained RelRe model to describe the replacement of lineages where some lineages have the same transmissibility. A constrained RelRe model, which is a constrained version of the original RelRe model, is obtained by introducing equality constraints among the *R*_*RI*_ of lineages in the original RelRe model. We can also consider such a constrained version of other lineage replacement models that use relative effective reproduction numbers. One of such models is the lineage replacement model that uses a softmax link function (22). By finding a constraining partition that minimizes the AIC of such replacement models using the FindPart-*w* algorithm, one can identify lineages that have the same transmissibility.

Using a hypothetical dataset, we observed that increasing the beam width *w* of the FindPart-*w* improved the AIC of the constrained RelRe models (Table 3). At the same time, however, increasing the beam width increased the number of maximum likelihood calculations (Figure 3). These results demonstrated the trade-off between computational time and achieved AIC. However, the difference in AIC was small enough to conclude that the constrained RelRe models found by FindPart-1 were as good as those found by FindPart-50 and FindPart-40 in terms of AIC. We also found that FindPart-1 and FindPart-20 were not significantly different from FindPart-50 in terms of the sensitivity, specificity, and precision (Table 4). Considering these observations, we can conclude that FindPart-1 is a reasonable choice among other *w*, when computational time is limited.

Apart from hierarchical clustering, the *k*-means algorithm (49) is another popular clustering algorithm. When the partitions found by FindPart-50 were compared with clustering results of the *k*-means using AIC, FindPart-*w* showed significantly better performances than *k*-means when *n* ≥ 30 (Table 3). However, when the partitions found by FindPart-50 and clustering results of the *k*-means were compared with the true partitions with which observation datasets were generated, *k*-means showed significantly better specificities than FindPart-50 (Table 4). A possible reason for this may be the existence of two lineages that have almost the same but slightly different *R*_*RI*_ in the hypothetical observation dataset. This may happen frequently because the true parameters for *R*_*RI*_ of multiple lineages were randomly generated from a uniform distribution. FindPart-*w* may have grouped such lineages having very similar *R*_*RI*_ together in order to minimise the AIC of constrained RelRe models in a more aggressive manner than the *k*-means algorithm. Therefore, FindPart-*w* may be more prone to have false positives, potentially leading to lower specificity compared to the *k*-means algorithm.

In this study, we applied the FindPart-*w* algorithm to Omicron sub-lineages of SARS-CoV-2. Naturally, FindPart-*w* can be applicable not only to SARS-CoV-2 but also to other viruses that have multiple lineages. To apply FindPart-*w* to another virus, it requires information on the generation time distribution of infections and abundant nucleotide sequences sampled from a single population, each of which is labelled with a lineage name and the time of sample collection. Influenza A virus is an example of a target virus to which the FindPart-*w* algorithm can be applied.

One interesting application of the constrained RelRe model found by FindPart-*w* is the investigation of the virological function of amino acid substitutions. FindPart-*w* can identify lineages that share the same *R*_*RI*_ and output results in the form of a partition of lineages. Two lineages in the same block in the output partition have the same transmissibility. This implies that combinations of amino acid substitutions found between two lineages in the same block do not alter the transmissibility of these lineages. In contrast, two lineages in different blocks in the output partition have different transmissibility. This implies that combinations of amino acid substitutions between two lineages in different blocks do alter the transmissibility of these lineages. Based on such information, one can investigate amino acid positions on a protein at which substitutions alter the transmissibility of a virus.

From actual observation data of lineage counts in the United States, the FindPart-1 algorithm identified groups of Pango lineages that share the same transmissibilities. Two lineages that were identified to share the same transmissibilities in this dataset may not share the same transmissibilities in other datasets from other countries or regions. This is because the immune level against each lineage in one population may be different from that in another population due to their different epidemic and vaccination histories. This is a limitation to consider when trying to use our method for developing future lineage designation systems. To solve this problem, we need to identify lineages or viruses having the same transmissibility using not only data from a single population but also data from multiple populations in the world.

FindPart-*w* clusters together two lineages by considering only their transmissibility, but it does not consider their phylogenetic relationships. For this reason, FindPart-*w* may group two lineages with different genetic backgrounds into the same block. To solve this problem, a mechanism that prevents the clustering of two genetically distant lineages needs to be introduced. A possible way to do this would be to use a similar approach to the DBSCAN algorithm (56). For example, we may restrict the FindPart-*w* algorithm so that two blocks are merged only when the genetic distance between a lineage in the first block and another lineage in the second block is smaller than or equal to a given threshold. The genetic distance between two lineages can be defined, for instance, as the number of amino acid substitutions between their consensus amino acid sequences of some proteins. The number of edges that connect two lineages on the Pango lineage tree can also be used as a genetic distance. From the nature of the Pango lineage system, the number of connecting edges between two lineages can be determined by comparing their unaliased Pango lineage names. Taking genetic backgrounds among lineages into account, we may extend the FindPart-*w* algorithm to identify groups of lineages that have the same transmissibility and similar genetic backgrounds.

## Conclusion

By developing the FindPart-*w* algorithm, this study showed that viruses that had the same transmissibility were able to be identified from time-stamped lineage counts of viral sequences in a practical computational time. The findings will contribute to the future development of lineage designation systems that consider both genetic backgrounds and transmissibilities of viral strains.

## Data Availability

The values used to build Figure 2, raw values used to get statistics in Table 1, Table 2, Table 3, and Table 4, the metadata of sequences downloaded from GISAID database can be found in the supplementary tables. The program code of the FindPart-*w* algorithm can be downloaded from our GitHub repository https://github.com/musonda-richard/FindPart-w/. The dataset of the actual sequence counts from the United States, which were used to create Table 5, Table 6, and Table 7, are included in the same GitHub repository.

## Author Contributions

Conceptualization: Kimihito I.; Data Curation: R.M.; Formal Analysis: R.M., Kimihito I.; Funding Acquisition: R.O., Kimihito I.; Investigation: R.M.; Methodology: Kimihito I.; Project Administration: Kimihito I.; Resources: R.O., Kimihito I.; Software: R.M., Kimihito I.; Supervision: Koichi I., Kimihito I.; Validation: Kimihito I.; Visualization: R.M.; Writing - Original Draft Preparation: R.M., K.I.; Writing - Review & Editing: R.O., Koichi I. All authors have read and agreed to the published version of the manuscript.

## Acknowledgments

We would like to acknowledge the funders of this project. The development of the programs used in this work was supported by the Japan Agency for Medical Research and Development (grant numbers JP24fk0108685, JP24wm0125008). Kimihito I. received funding for JSPS KAKENHI (21H03490). R.M. was supported by the Ministry of Education, Culture, Sports, Science, and Technology (MEXT) scholarship, from the Government of Japan. The funders had no role of influence in the study design, data collection and analysis, decision to publish, or drafting and preparation of the manuscript.

We wish to give thanks, gratitude and acknowledgment to the laboratories and personnel responsible for obtaining the specimens and the laboratories where genetic sequence data were generated and shared through the GISAID Initiative. Without the work of these people this research might not have been possible. The details on originating laboratories, submitting laboratories, and authors of SARS-CoV-2 sequence data can be found in Table S4.

