## Supplemental Materials for "Identifying SARS-CoV-2 Lineages that Share the Same Relative Effective Reproduction Numbers"

**Table S1. The details on originating laboratories, submitting laboratories, and authors of SARS-CoV-2 sequence data**

The table is provided as a separate Excel file in the supplementary material.

**Table S2. Raw values of the number of maximum likelihood estimations required by the Naïve, FindPart-**$\boldsymbol{w (w=\infty, 1, 10, 20,30,40,50)}$**,** $\boldsymbol{k}$**-means, and hierarchical clustering algorithms.**

| n | run | naive | $w$=$\infty$ | $w$=1 | $w$=10 | $w$=20 | $w$=30 | $w$=40 | $w$=50 | $k$-means | hierarchical |
| --- | --- | --- | --- | --- | --- | --- | --- | --- | --- | --- | --- |
| 5 | 1 | 52 | 12 | 12 | 12 | 12 | 12 | 12 | 12 | 25 | 21 |
| 5 | 2 | 52 | 12 | 12 | 12 | 12 | 12 | 12 | 12 | 25 | 21 |
| 5 | 3 | 52 | 15 | 13 | 15 | 15 | 15 | 15 | 15 | 25 | 21 |
| 5 | 4 | 52 | 12 | 12 | 12 | 12 | 12 | 12 | 12 | 25 | 21 |
| 5 | 5 | 52 | 12 | 12 | 12 | 12 | 12 | 12 | 12 | 25 | 21 |
| 5 | 6 | 52 | 12 | 12 | 12 | 12 | 12 | 12 | 12 | 25 | 21 |
| 5 | 7 | 52 | 12 | 12 | 12 | 12 | 12 | 12 | 12 | 25 | 21 |
| 5 | 8 | 52 | 12 | 12 | 12 | 12 | 12 | 12 | 12 | 25 | 21 |
| 5 | 9 | 52 | 11 | 11 | 11 | 11 | 11 | 11 | 11 | 25 | 21 |
| 5 | 10 | 52 | 12 | 12 | 12 | 12 | 12 | 12 | 12 | 25 | 21 |
| 10 | 1 | 115975 | 50 | 49 | 50 | 50 | 50 | 50 | 50 | 100 | 166 |
| 10 | 2 | 115975 | 50 | 49 | 50 | 50 | 50 | 50 | 50 | 100 | 166 |
| 10 | 3 | 115975 | 60 | 53 | 60 | 60 | 60 | 60 | 60 | 100 | 166 |
| 10 | 4 | 115975 | 74 | 55 | 74 | 74 | 74 | 74 | 74 | 100 | 166 |
| 10 | 5 | 115975 | 223 | 66 | 174 | 212 | 220 | 222 | 225 | 100 | 166 |
| 10 | 6 | 115975 | 69 | 53 | 69 | 69 | 69 | 69 | 69 | 100 | 166 |
| 10 | 7 | 115975 | 64 | 55 | 64 | 64 | 64 | 64 | 64 | 100 | 166 |
| 10 | 8 | 115975 | 254 | 67 | 183 | 224 | 247 | 252 | 253 | 100 | 166 |
| 10 | 9 | 115975 | 70 | 56 | 70 | 70 | 70 | 70 | 70 | 100 | 166 |
| 10 | 10 | 115975 | 82 | 56 | 82 | 82 | 82 | 82 | 82 | 100 | 166 |
| 15 | 1 | 1382958545 | 842 | 141 | 367 | 501 | 591 | 669 | 733 | 225 | 561 |
| 15 | 2 | 1382958545 | 514 | 145 | 359 | 458 | 503 | 514 | 514 | 225 | 561 |
| 15 | 3 | 1382958545 | 453 | 139 | 297 | 366 | 412 | 440 | 452 | 225 | 561 |
| 15 | 4 | 1382958545 | 49198 | 227 | 1159 | 2051 | 2786 | 3422 | 4006 | 225 | 561 |
| 15 | 5 | 1382958545 | 476 | 138 | 322 | 414 | 444 | 463 | 475 | 225 | 561 |
| 15 | 6 | 1382958545 | 834 | 155 | 410 | 559 | 660 | 718 | 756 | 225 | 561 |
| 15 | 7 | 1382958545 | 389 | 136 | 288 | 351 | 379 | 385 | 388 | 225 | 561 |
| 15 | 8 | 1382958545 | 3036 | 165 | 647 | 1014 | 1268 | 1470 | 1688 | 225 | 561 |
| 15 | 9 | 1382958545 | 414 | 145 | 344 | 412 | 410 | 410 | 413 | 225 | 561 |
| 15 | 10 | 1382958545 | 802 | 144 | 361 | 476 | 549 | 610 | 650 | 225 | 561 |
| 20 | 1 | $5.17\times{10}^{13}$ | 5908 | 286 | 941 | 1464 | 1869 | 2227 | 2553 | 400 | 1331 |
| 20 | 2 | $5.17\times{10}^{13}$ | 5670 | 255 | 714 | 1075 | 1363 | 1642 | 1897 | 400 | 1331 |
| 20 | 3 | $5.17\times{10}^{13}$ | 58044 | 344 | 1420 | 2407 | 3290 | 4109 | 4847 | 400 | 1331 |
| 20 | 4 | $5.17\times{10}^{13}$ | 14077 | 286 | 924 | 1488 | 1927 | 2347 | 2727 | 400 | 1331 |
| 20 | 5 | $5.17\times{10}^{13}$ | 37803 | 300 | 1165 | 1964 | 2645 | 3219 | 3741 | 400 | 1331 |
| 20 | 6 | $5.17\times{10}^{13}$ | 39628 | 286 | 978 | 1594 | 2117 | 2566 | 2988 | 400 | 1331 |
| 20 | 7 | $5.17\times{10}^{13}$ | 22301 | 309 | 1217 | 2075 | 2789 | 3472 | 4108 | 400 | 1331 |
| 20 | 8 | $5.17\times{10}^{13}$ | 53332 | 316 | 1283 | 2197 | 2915 | 3602 | 4241 | 400 | 1331 |
| 20 | 9 | $5.17\times{10}^{13}$ | 4457 | 253 | 702 | 1057 | 1341 | 1618 | 1842 | 400 | 1331 |
| 20 | 10 | $5.17\times{10}^{13}$ | 46124 | 353 | 1470 | 2580 | 3530 | 4329 | 5112 | 400 | 1331 |
| 25 | 1 | $4.64\times{10}^{18}$ | n.a. | 521 | 2321 | 4073 | 5709 | 7204 | 8576 | 625 | 2601 |
| 25 | 2 | $4.64\times{10}^{18}$ | n.a. | 784 | 4918 | 9338 | 13647 | 17872 | 21954 | 625 | 2601 |
| 25 | 3 | $4.64\times{10}^{18}$ | n.a. | 471 | 1728 | 2952 | 3989 | 4916 | 5814 | 625 | 2601 |
| 25 | 4 | $4.64\times{10}^{18}$ | n.a. | 594 | 3112 | 5804 | 8231 | 10702 | 12977 | 625 | 2601 |
| 25 | 5 | $4.64\times{10}^{18}$ | n.a. | 648 | 3758 | 7106 | 10244 | 13241 | 16175 | 625 | 2601 |
| 25 | 6 | $4.64\times{10}^{18}$ | n.a. | 528 | 2412 | 4232 | 5887 | 7453 | 8898 | 625 | 2601 |
| 25 | 7 | $4.64\times{10}^{18}$ | n.a. | 587 | 3160 | 5763 | 8283 | 10550 | 12766 | 625 | 2601 |
| 25 | 8 | $4.64\times{10}^{18}$ | n.a. | 909 | 6001 | 11173 | 16099 | 20868 | 25454 | 625 | 2601 |
| 25 | 9 | $4.64\times{10}^{18}$ | n.a. | 841 | 5207 | 9973 | 14602 | 19043 | 23363 | 625 | 2601 |
| 25 | 10 | $4.64\times{10}^{18}$ | n.a. | 521 | 2254 | 4078 | 5764 | 7282 | 8797 | 625 | 2601 |
| 30 | 1 | $8.47\times{10}^{23}$ | n.a. | 693 | 2906 | 5146 | 7189 | 9097 | 10888 | 900 | 4496 |
| 30 | 2 | $8.47\times{10}^{23}$ | n.a. | 837 | 4543 | 8304 | 11894 | 15361 | 18623 | 900 | 4496 |
| 30 | 3 | $8.47\times{10}^{23}$ | n.a. | 842 | 4279 | 8046 | 11699 | 15114 | 18554 | 900 | 4496 |
| 30 | 4 | $8.47\times{10}^{23}$ | n.a. | 883 | 4364 | 7967 | 11448 | 14829 | 18030 | 900 | 4496 |
| 30 | 5 | $8.47\times{10}^{23}$ | n.a. | 795 | 3808 | 6939 | 9934 | 12742 | 15526 | 900 | 4496 |
| 30 | 6 | $8.47\times{10}^{23}$ | n.a. | 865 | 4449 | 8210 | 11768 | 15322 | 18718 | 900 | 4496 |
| 30 | 7 | $8.47\times{10}^{23}$ | n.a. | 807 | 3913 | 7349 | 10385 | 13402 | 16198 | 900 | 4496 |
| 30 | 8 | $8.47\times{10}^{23}$ | n.a. | 883 | 4280 | 8068 | 11651 | 15085 | 18385 | 900 | 4496 |
| 30 | 9 | $8.47\times{10}^{23}$ | n.a. | 1051 | 6427 | 12185 | 17664 | 22970 | 28033 | 900 | 4496 |
| 30 | 10 | $8.47\times{10}^{23}$ | n.a. | 1098 | 6779 | 13026 | 19229 | 25300 | 31117 | 900 | 4496 |
| 35 | 1 | $2.82\times{10}^{29}$ | n.a. | 1182 | 6344 | 11862 | 17142 | 22283 | 27412 | 1225 | 7141 |
| 35 | 2 | $2.82\times{10}^{29}$ | n.a. | 1321 | 7240 | 13855 | 20342 | 26376 | 32304 | 1225 | 7141 |
| 35 | 3 | $2.82\times{10}^{29}$ | n.a. | 1280 | 6749 | 12309 | 17842 | 23336 | 28469 | 1225 | 7141 |
| 35 | 4 | $2.82\times{10}^{29}$ | n.a. | 1414 | 8373 | 16029 | 23303 | 30311 | 37255 | 1225 | 7141 |
| 35 | 5 | $2.82\times{10}^{29}$ | n.a. | 1452 | 8459 | 16099 | 23648 | 31162 | 38440 | 1225 | 7141 |
| 35 | 6 | $2.82\times{10}^{29}$ | n.a. | 1753 | 12385 | 24473 | 35798 | 46959 | 57711 | 1225 | 7141 |
| 35 | 7 | $2.82\times{10}^{29}$ | n.a. | 1364 | 8205 | 15367 | 22575 | 29482 | 36385 | 1225 | 7141 |
| 35 | 8 | $2.82\times{10}^{29}$ | n.a. | 1224 | 6510 | 12101 | 17541 | 22870 | 28049 | 1225 | 7141 |
| 35 | 9 | $2.82\times{10}^{29}$ | n.a. | 1594 | 9991 | 19290 | 28295 | 37491 | 46267 | 1225 | 7141 |
| 35 | 10 | $2.82\times{10}^{29}$ | n.a. | 1217 | 6514 | 12548 | 18104 | 23559 | 29128 | 1225 | 7141 |

*A cell with n.a. indicates missing values where we were unable to finish the calculation due to long computational time.

**Table S3. Raw AIC values of partitions output by the original RelRe model, FindPart-**$\boldsymbol{w (w=\infty,1,10,20,30,40,50)}$**, and** $\boldsymbol{k}$**-means clustering**

| Procedure to find the best partition |  | Number of lineages (n) | | | | | | |  |  |
| --- | --- | --- | --- | --- | --- | --- | --- | --- | --- | --- |
|  | run | Original RelRe | $w=\infty$ | $w=1$ | $w=10$ | $w=20$ | $w=30$ | $w=40$ | $w=50$ | $k$-means |
| 5 | 1 | 1830.89 | 1827.45 | 1827.45 | 1827.45 | 1827.45 | 1827.45 | 1827.45 | 1827.45 | 1827.45 |
|  | 2 | 2022.61 | 2018.71 | 2018.71 | 2018.71 | 2018.71 | 2018.71 | 2018.71 | 2018.71 | 2018.71 |
|  | 3 | 1983.36 | 1981.85 | 1981.85 | 1981.85 | 1981.85 | 1981.85 | 1981.85 | 1981.85 | 1981.85 |
|  | 4 | 2178.15 | 2174.93 | 2174.93 | 2174.93 | 2174.93 | 2174.93 | 2174.93 | 2174.93 | 2174.93 |
|  | 5 | 2089.95 | 2085.96 | 2085.96 | 2085.96 | 2085.96 | 2085.96 | 2085.96 | 2085.96 | 2085.96 |
|  | 6 | 1498.81 | 1494.61 | 1494.61 | 1494.61 | 1494.61 | 1494.61 | 1494.61 | 1494.61 | 1494.61 |
|  | 7 | 1530.03 | 1526.28 | 1526.28 | 1526.28 | 1526.28 | 1526.28 | 1526.28 | 1526.28 | 1526.28 |
|  | 8 | 1841.67 | 1837.98 | 1837.98 | 1837.98 | 1837.98 | 1837.98 | 1837.98 | 1837.98 | 1837.98 |
|  | 9 | 1264.59 | 1262.58 | 1262.58 | 1262.58 | 1262.58 | 1262.58 | 1262.58 | 1262.58 | 1262.58 |
|  | 10 | 2174.63 | 2171.37 | 2171.37 | 2171.37 | 2171.37 | 2171.37 | 2171.37 | 2171.37 | 2171.37 |
| 10 | 1 | 4098.75 | 4094.08 | 4094.08 | 4094.08 | 4094.08 | 4094.08 | 4094.08 | 4094.08 | 4094.08 |
|  | 2 | 4632.74 | 4628.19 | 4628.19 | 4628.19 | 4628.19 | 4628.19 | 4628.19 | 4628.19 | 4628.19 |
|  | 3 | 4118.06 | 4113.25 | 4113.25 | 4113.25 | 4113.25 | 4113.25 | 4113.25 | 4113.25 | 4113.25 |
|  | 4 | 4096.62 | 4091.92 | 4091.92 | 4091.92 | 4091.92 | 4091.92 | 4091.92 | 4091.92 | 4091.92 |
|  | 5 | 2711.35 | 2705.11 | 2705.11 | 2705.11 | 2705.11 | 2705.11 | 2705.11 | 2705.11 | 2705.11 |
|  | 6 | 3400.96 | 3393.35 | 3393.35 | 3393.35 | 3393.35 | 3393.35 | 3393.35 | 3393.35 | 3393.35 |
|  | 7 | 3840.24 | 3833.12 | 3833.12 | 3833.12 | 3833.12 | 3833.12 | 3833.12 | 3833.12 | 3833.12 |
|  | 8 | 3547.14 | 3541.75 | 3541.75 | 3541.75 | 3541.75 | 3541.75 | 3541.75 | 3541.75 | 3542.71 |
|  | 9 | 3178.62 | 3173.24 | 3173.24 | 3173.24 | 3173.24 | 3173.24 | 3173.24 | 3173.24 | 3173.24 |
|  | 10 | 4749.86 | 4745.96 | 4745.96 | 4745.96 | 4745.96 | 4745.96 | 4745.96 | 4745.96 | 4745.96 |
| 15 | 1 | 7146.75 | 7136.19 | 7136.19 | 7136.19 | 7136.19 | 7136.19 | 7136.19 | 7136.19 | 7136.19 |
|  | 2 | 6042.12 | 6039.24 | 6039.24 | 6039.24 | 6039.24 | 6039.24 | 6039.24 | 6039.24 | 6039.28 |
|  | 3 | 5697.25 | 5686.35 | 5686.35 | 5686.35 | 5686.35 | 5686.35 | 5686.35 | 5686.35 | 5686.35 |
|  | 4 | 3507.48 | 3499.74 | 3499.74 | 3499.74 | 3499.74 | 3499.74 | 3499.74 | 3499.74 | 3499.74 |
|  | 5 | 5874.01 | 5868.11 | 5868.11 | 5868.11 | 5868.11 | 5868.11 | 5868.11 | 5868.11 | 5871.33 |
|  | 6 | 5551.92 | 5541.94 | 5541.94 | 5541.94 | 5541.94 | 5541.94 | 5541.94 | 5541.94 | 5541.94 |
|  | 7 | 7153.42 | 7144.72 | 7144.72 | 7144.72 | 7144.72 | 7144.72 | 7144.72 | 7144.72 | 7144.72 |
|  | 8 | 5512.48 | 5499.82 | 5499.82 | 5499.82 | 5499.82 | 5499.82 | 5499.82 | 5499.82 | 5499.82 |
|  | 9 | 6305.97 | 6296.79 | 6298.26 | 6296.79 | 6296.79 | 6296.79 | 6296.79 | 6296.79 | 6298.21 |
|  | 10 | 6663.74 | 6655.19 | 6655.19 | 6655.19 | 6655.19 | 6655.19 | 6655.19 | 6655.19 | 6655.26 |
| 20 | 1 | 9079.98 | 9064.00 | 9064.00 | 9064.00 | 9064.00 | 9064.00 | 9064.00 | 9064.00 | 9064.00 |
|  | 2 | 10210.39 | 10194.46 | 10194.46 | 10194.46 | 10194.46 | 10194.46 | 10194.46 | 10194.46 | 10194.46 |
|  | 3 | 8411.76 | 8398.67 | 8398.67 | 8398.67 | 8398.67 | 8398.67 | 8398.67 | 8398.67 | 8398.67 |
|  | 4 | 7817.15 | 7799.39 | 7799.39 | 7799.39 | 7799.39 | 7799.39 | 7799.39 | 7799.39 | 7799.39 |
|  | 5 | 7829.43 | 7809.23 | 7809.23 | 7809.23 | 7809.23 | 7809.23 | 7809.23 | 7809.23 | 7809.23 |
|  | 6 | 8286.51 | 8268.97 | 8268.97 | 8268.97 | 8268.97 | 8268.97 | 8268.97 | 8268.97 | 8268.97 |
|  | 7 | 8142.28 | 8125.31 | 8125.31 | 8125.31 | 8125.31 | 8125.31 | 8125.31 | 8125.31 | 8125.31 |
|  | 8 | 8107.76 | 8090.82 | 8090.82 | 8090.82 | 8090.82 | 8090.82 | 8090.82 | 8090.82 | 8090.82 |
|  | 9 | 8801.20 | 8790.03 | 8790.03 | 8790.03 | 8790.03 | 8790.03 | 8790.03 | 8790.03 | 8795.53 |
|  | 10 | 8066.08 | 8043.61 | 8043.61 | 8043.61 | 8043.61 | 8043.61 | 8043.61 | 8043.61 | 8043.61 |
| 25 | 1 | 9798.64 | n.a.* | 9776.93 | 9776.93 | 9776.93 | 9776.93 | 9776.93 | 9776.93 | 9780.19 |
|  | 2 | 8334.80 | n.a. | 8312.35 | 8309.86 | 8309.95 | 8309.86 | 8309.86 | 8309.86 | 8310.83 |
|  | 3 | 11002.11 | n.a. | 10982.82 | 10982.82 | 10982.82 | 10982.82 | 10982.82 | 10982.82 | 10983.70 |
|  | 4 | 10532.03 | n.a. | 10500.37 | 10500.37 | 10500.37 | 10500.37 | 10500.37 | 10500.37 | 10502.30 |
|  | 5 | 9028.25 | n.a. | 9001.72 | 9001.00 | 9001.00 | 9001.00 | 9001.00 | 9001.00 | 9007.31 |
|  | 6 | 11612.93 | n.a. | 11591.52 | 11591.52 | 11591.52 | 11591.52 | 11591.52 | 11591.52 | 11593.96 |
|  | 7 | 10191.57 | n.a. | 10168.77 | 10168.77 | 10167.90 | 10167.88 | 10167.88 | 10167.88 | 10171.31 |
|  | 8 | 8237.42 | n.a. | 8209.86 | 8209.84 | 8209.84 | 8209.84 | 8209.84 | 8209.84 | 8209.84 |
|  | 9 | 8300.77 | n.a. | 8277.41 | 8275.40 | 8275.40 | 8275.40 | 8275.40 | 8275.40 | 8275.75 |
|  | 10 | 10032.10 | n.a. | 10020.77 | 10020.02 | 10020.02 | 10020.02 | 10020.02 | 10020.02 | 10020.02 |
| 30 | 1 | 15588.84 | n.a. | 15557.66 | 15556.99 | 15556.99 | 15556.99 | 15556.99 | 15556.99 | 15558.71 |
|  | 2 | 13320.74 | n.a. | 13288.05 | 13288.05 | 13288.05 | 13288.05 | 13288.05 | 13288.05 | 13288.68 |
|  | 3 | 13136.84 | n.a. | 13107.83 | 13107.32 | 13107.32 | 13107.32 | 13107.32 | 13107.32 | 13108.45 |
|  | 4 | 14008.20 | n.a. | 13982.40 | 13982.40 | 13982.40 | 13982.40 | 13982.40 | 13982.40 | 13982.40 |
|  | 5 | 14210.79 | n.a. | 14184.90 | 14184.90 | 14184.90 | 14184.90 | 14184.90 | 14184.90 | 14184.90 |
|  | 6 | 13712.93 | n.a. | 13681.88 | 13681.88 | 13681.88 | 13681.88 | 13681.88 | 13681.88 | 13689.03 |
|  | 7 | 14448.23 | n.a. | 14421.48 | 14421.00 | 14421.00 | 14421.00 | 14421.00 | 14421.00 | 14423.80 |
|  | 8 | 11936.12 | n.a. | 11904.67 | 11904.67 | 11904.67 | 11904.67 | 11904.67 | 11904.67 | 11904.67 |
|  | 9 | 10584.93 | n.a. | 10553.36 | 10544.89 | 10543.53 | 10543.53 | 10543.53 | 10543.53 | 10553.81 |
|  | 10 | 12039.36 | n.a. | 12010.93 | 12010.68 | 12010.68 | 12010.68 | 12010.68 | 12010.68 | 12010.68 |
| 35 | 1 | 15575.84 | n.a. | 15545.39 | 15545.39 | 15545.39 | 15545.39 | 15545.39 | 15545.39 | 15547.75 |
|  | 2 | 14478.62 | n.a. | 14422.93 | 14422.93 | 14422.93 | 14422.93 | 14422.93 | 14422.93 | 14423.43 |
|  | 3 | 15791.32 | n.a. | 15761.30 | 15760.24 | 15759.49 | 15759.49 | 15759.49 | 15759.49 | 15763.43 |
|  | 4 | 14040.79 | n.a. | 13976.29 | 13975.60 | 13974.65 | 13974.65 | 13974.65 | 13974.65 | 13982.66 |
|  | 5 | 15532.37 | n.a. | 15488.65 | 15487.56 | 15487.56 | 15487.56 | 15487.56 | 15487.56 | 15487.56 |
|  | 6 | 13070.63 | n.a. | 13010.10 | 13010.18 | 13009.56 | 13009.53 | 13009.53 | 13009.53 | 13015.60 |
|  | 7 | 14266.28 | n.a. | 14219.07 | 14217.79 | 14217.79 | 14217.79 | 14217.79 | 14217.79 | 14223.85 |
|  | 8 | 16407.29 | n.a. | 16381.70 | 16381.37 | 16381.37 | 16381.37 | 16381.37 | 16381.37 | 16381.70 |
|  | 9 | 13537.93 | n.a. | 13485.53 | 13485.35 | 13485.35 | 13485.35 | 13484.83 | 13484.83 | 13487.25 |
|  | 10 | 15506.07 | n.a. | 15469.30 | 15468.44 | 15468.44 | 15468.44 | 15468.44 | 15468.44 | 15470.54 |

*A cell with n.a. indicates missing values where we were unable to finish the calculation due to long computational time.

**Table S4. Raw values of sensitivity, specificity, and precision**

| n | run | N | algorithm | precision | sensitivity | specificity |
| --- | --- | --- | --- | --- | --- | --- |
| 5 | 1 | 500 | FindPart-1 | 1.000 | 1.000 | 1.000 |
| 5 | 1 | 500 | FindPart-20 | 1.000 | 1.000 | 1.000 |
| 5 | 1 | 500 | FindPart-50 | 1.000 | 1.000 | 1.000 |
| 5 | 1 | 500 | k-means | 1.000 | 1.000 | 1.000 |
| 5 | 2 | 500 | FindPart-1 | 1.000 | 1.000 | 1.000 |
| 5 | 2 | 500 | FindPart-20 | 1.000 | 1.000 | 1.000 |
| 5 | 2 | 500 | FindPart-50 | 1.000 | 1.000 | 1.000 |
| 5 | 2 | 500 | k-means | 1.000 | 1.000 | 1.000 |
| 5 | 3 | 500 | FindPart-1 | 1.000 | 1.000 | 1.000 |
| 5 | 3 | 500 | FindPart-20 | 1.000 | 1.000 | 1.000 |
| 5 | 3 | 500 | FindPart-50 | 1.000 | 1.000 | 1.000 |
| 5 | 3 | 500 | k-means | 1.000 | 1.000 | 1.000 |
| 5 | 4 | 500 | FindPart-1 | 1.000 | 1.000 | 1.000 |
| 5 | 4 | 500 | FindPart-20 | 1.000 | 1.000 | 1.000 |
| 5 | 4 | 500 | FindPart-50 | 1.000 | 1.000 | 1.000 |
| 5 | 4 | 500 | k-means | 1.000 | 1.000 | 1.000 |
| 5 | 5 | 500 | FindPart-1 | 1.000 | 1.000 | 1.000 |
| 5 | 5 | 500 | FindPart-20 | 1.000 | 1.000 | 1.000 |
| 5 | 5 | 500 | FindPart-50 | 1.000 | 1.000 | 1.000 |
| 5 | 5 | 500 | k-means | 1.000 | 1.000 | 1.000 |
| 5 | 6 | 500 | FindPart-1 | 1.000 | 1.000 | 1.000 |
| 5 | 6 | 500 | FindPart-20 | 1.000 | 1.000 | 1.000 |
| 5 | 6 | 500 | FindPart-50 | 1.000 | 1.000 | 1.000 |
| 5 | 6 | 500 | k-means | 1.000 | 1.000 | 1.000 |
| 5 | 7 | 500 | FindPart-1 | 1.000 | 1.000 | 1.000 |
| 5 | 7 | 500 | FindPart-20 | 1.000 | 1.000 | 1.000 |
| 5 | 7 | 500 | FindPart-50 | 1.000 | 1.000 | 1.000 |
| 5 | 7 | 500 | k-means | 1.000 | 1.000 | 1.000 |
| 5 | 8 | 500 | FindPart-1 | 1.000 | 1.000 | 1.000 |
| 5 | 8 | 500 | FindPart-20 | 1.000 | 1.000 | 1.000 |
| 5 | 8 | 500 | FindPart-50 | 1.000 | 1.000 | 1.000 |
| 5 | 8 | 500 | k-means | 1.000 | 1.000 | 1.000 |
| 5 | 9 | 500 | FindPart-1 | 1.000 | 0.500 | 1.000 |
| 5 | 9 | 500 | FindPart-20 | 1.000 | 0.500 | 1.000 |
| 5 | 9 | 500 | FindPart-50 | 1.000 | 0.500 | 1.000 |
| 5 | 9 | 500 | k-means | 1.000 | 0.500 | 1.000 |
| 5 | 10 | 500 | FindPart-1 | 1.000 | 1.000 | 1.000 |
| 5 | 10 | 500 | FindPart-20 | 1.000 | 1.000 | 1.000 |
| 5 | 10 | 500 | FindPart-50 | 1.000 | 1.000 | 1.000 |
| 5 | 10 | 500 | k-means | 1.000 | 1.000 | 1.000 |
| 10 | 1 | 1000 | FindPart-1 | 1.000 | 0.667 | 1.000 |
| 10 | 1 | 1000 | FindPart-20 | 1.000 | 0.667 | 1.000 |
| 10 | 1 | 1000 | FindPart-50 | 1.000 | 0.667 | 1.000 |
| 10 | 1 | 1000 | k-means | 1.000 | 0.667 | 1.000 |
| 10 | 2 | 1000 | FindPart-1 | 1.000 | 1.000 | 1.000 |
| 10 | 2 | 1000 | FindPart-20 | 1.000 | 1.000 | 1.000 |
| 10 | 2 | 1000 | FindPart-50 | 1.000 | 1.000 | 1.000 |
| 10 | 2 | 1000 | k-means | 1.000 | 1.000 | 1.000 |
| 10 | 3 | 1000 | FindPart-1 | 1.000 | 1.000 | 1.000 |
| 10 | 3 | 1000 | FindPart-20 | 1.000 | 1.000 | 1.000 |
| 10 | 3 | 1000 | FindPart-50 | 1.000 | 1.000 | 1.000 |
| 10 | 3 | 1000 | k-means | 1.000 | 1.000 | 1.000 |
| 10 | 4 | 1000 | FindPart-1 | 1.000 | 1.000 | 1.000 |
| 10 | 4 | 1000 | FindPart-20 | 1.000 | 1.000 | 1.000 |
| 10 | 4 | 1000 | FindPart-50 | 1.000 | 1.000 | 1.000 |
| 10 | 4 | 1000 | k-means | 1.000 | 1.000 | 1.000 |
| 10 | 5 | 1000 | FindPart-1 | 0.800 | 1.000 | 0.976 |
| 10 | 5 | 1000 | FindPart-20 | 0.800 | 1.000 | 0.976 |
| 10 | 5 | 1000 | FindPart-50 | 0.800 | 1.000 | 0.976 |
| 10 | 5 | 1000 | k-means | 0.800 | 1.000 | 0.976 |
| 10 | 6 | 1000 | FindPart-1 | 0.667 | 0.667 | 0.976 |
| 10 | 6 | 1000 | FindPart-20 | 0.667 | 0.667 | 0.976 |
| 10 | 6 | 1000 | FindPart-50 | 0.667 | 0.667 | 0.976 |
| 10 | 6 | 1000 | k-means | 0.667 | 0.667 | 0.976 |
| 10 | 7 | 1000 | FindPart-1 | 0.750 | 1.000 | 0.976 |
| 10 | 7 | 1000 | FindPart-20 | 0.750 | 1.000 | 0.976 |
| 10 | 7 | 1000 | FindPart-50 | 0.750 | 1.000 | 0.976 |
| 10 | 7 | 1000 | k-means | 0.750 | 1.000 | 0.976 |
| 10 | 8 | 1000 | FindPart-1 | 0.250 | 0.250 | 0.927 |
| 10 | 8 | 1000 | FindPart-20 | 0.250 | 0.250 | 0.927 |
| 10 | 8 | 1000 | FindPart-50 | 0.250 | 0.250 | 0.927 |
| 10 | 8 | 1000 | k-means | 0.400 | 0.500 | 0.927 |
| 10 | 9 | 1000 | FindPart-1 | 0.667 | 0.667 | 0.976 |
| 10 | 9 | 1000 | FindPart-20 | 0.667 | 0.667 | 0.976 |
| 10 | 9 | 1000 | FindPart-50 | 0.667 | 0.667 | 0.976 |
| 10 | 9 | 1000 | k-means | 0.667 | 0.667 | 0.976 |
| 10 | 10 | 1000 | FindPart-1 | 1.000 | 1.000 | 1.000 |
| 10 | 10 | 1000 | FindPart-20 | 1.000 | 1.000 | 1.000 |
| 10 | 10 | 1000 | FindPart-50 | 1.000 | 1.000 | 1.000 |
| 10 | 10 | 1000 | k-means | 1.000 | 1.000 | 1.000 |
| 15 | 1 | 1500 | FindPart-1 | 0.462 | 1.000 | 0.929 |
| 15 | 1 | 1500 | FindPart-20 | 0.462 | 1.000 | 0.929 |
| 15 | 1 | 1500 | FindPart-50 | 0.462 | 1.000 | 0.929 |
| 15 | 1 | 1500 | k-means | 0.462 | 1.000 | 0.929 |
| 15 | 2 | 1500 | FindPart-1 | 1.000 | 0.429 | 1.000 |
| 15 | 2 | 1500 | FindPart-20 | 1.000 | 0.429 | 1.000 |
| 15 | 2 | 1500 | FindPart-50 | 1.000 | 0.429 | 1.000 |
| 15 | 2 | 1500 | k-means | 1.000 | 0.714 | 1.000 |
| 15 | 3 | 1500 | FindPart-1 | 0.444 | 0.800 | 0.950 |
| 15 | 3 | 1500 | FindPart-20 | 0.444 | 0.800 | 0.950 |
| 15 | 3 | 1500 | FindPart-50 | 0.444 | 0.800 | 0.950 |
| 15 | 3 | 1500 | k-means | 0.444 | 0.800 | 0.950 |
| 15 | 4 | 1500 | FindPart-1 | 0.250 | 0.125 | 0.969 |
| 15 | 4 | 1500 | FindPart-20 | 0.250 | 0.125 | 0.969 |
| 15 | 4 | 1500 | FindPart-50 | 0.250 | 0.125 | 0.969 |
| 15 | 4 | 1500 | k-means | 0.250 | 0.125 | 0.969 |
| 15 | 5 | 1500 | FindPart-1 | 0.800 | 0.667 | 0.990 |
| 15 | 5 | 1500 | FindPart-20 | 0.800 | 0.667 | 0.990 |
| 15 | 5 | 1500 | FindPart-50 | 0.800 | 0.667 | 0.990 |
| 15 | 5 | 1500 | k-means | 0.667 | 0.333 | 0.990 |
| 15 | 6 | 1500 | FindPart-1 | 0.600 | 0.429 | 0.980 |
| 15 | 6 | 1500 | FindPart-20 | 0.600 | 0.429 | 0.980 |
| 15 | 6 | 1500 | FindPart-50 | 0.600 | 0.429 | 0.980 |
| 15 | 6 | 1500 | k-means | 0.600 | 0.429 | 0.980 |
| 15 | 7 | 1500 | FindPart-1 | 0.500 | 0.500 | 0.970 |
| 15 | 7 | 1500 | FindPart-20 | 0.500 | 0.500 | 0.970 |
| 15 | 7 | 1500 | FindPart-50 | 0.500 | 0.500 | 0.970 |
| 15 | 7 | 1500 | k-means | 0.500 | 0.500 | 0.970 |
| 15 | 8 | 1500 | FindPart-1 | 0.667 | 0.667 | 0.980 |
| 15 | 8 | 1500 | FindPart-20 | 0.667 | 0.667 | 0.980 |
| 15 | 8 | 1500 | FindPart-50 | 0.667 | 0.667 | 0.980 |
| 15 | 8 | 1500 | k-means | 0.667 | 0.667 | 0.980 |
| 15 | 9 | 1500 | FindPart-1 | 0.750 | 0.600 | 0.990 |
| 15 | 9 | 1500 | FindPart-20 | 1.000 | 1.000 | 1.000 |
| 15 | 9 | 1500 | FindPart-50 | 1.000 | 1.000 | 1.000 |
| 15 | 9 | 1500 | k-means | 1.000 | 0.800 | 1.000 |
| 15 | 10 | 1500 | FindPart-1 | 0.833 | 0.833 | 0.990 |
| 15 | 10 | 1500 | FindPart-20 | 0.833 | 0.833 | 0.990 |
| 15 | 10 | 1500 | FindPart-50 | 0.833 | 0.833 | 0.990 |
| 15 | 10 | 1500 | k-means | 0.833 | 0.833 | 0.990 |
| 20 | 1 | 2000 | FindPart-1 | 0.778 | 0.700 | 0.989 |
| 20 | 1 | 2000 | FindPart-20 | 0.778 | 0.700 | 0.989 |
| 20 | 1 | 2000 | FindPart-50 | 0.778 | 0.700 | 0.989 |
| 20 | 1 | 2000 | k-means | 0.778 | 0.700 | 0.989 |
| 20 | 2 | 2000 | FindPart-1 | 0.500 | 0.778 | 0.961 |
| 20 | 2 | 2000 | FindPart-20 | 0.500 | 0.778 | 0.961 |
| 20 | 2 | 2000 | FindPart-50 | 0.500 | 0.778 | 0.961 |
| 20 | 2 | 2000 | k-means | 0.500 | 0.778 | 0.961 |
| 20 | 3 | 2000 | FindPart-1 | 0.818 | 1.000 | 0.989 |
| 20 | 3 | 2000 | FindPart-20 | 0.818 | 1.000 | 0.989 |
| 20 | 3 | 2000 | FindPart-50 | 0.818 | 1.000 | 0.989 |
| 20 | 3 | 2000 | k-means | 0.818 | 1.000 | 0.989 |
| 20 | 4 | 2000 | FindPart-1 | 0.500 | 0.667 | 0.967 |
| 20 | 4 | 2000 | FindPart-20 | 0.500 | 0.667 | 0.967 |
| 20 | 4 | 2000 | FindPart-50 | 0.500 | 0.667 | 0.967 |
| 20 | 4 | 2000 | k-means | 0.500 | 0.667 | 0.967 |
| 20 | 5 | 2000 | FindPart-1 | 0.667 | 0.444 | 0.989 |
| 20 | 5 | 2000 | FindPart-20 | 0.667 | 0.444 | 0.989 |
| 20 | 5 | 2000 | FindPart-50 | 0.667 | 0.444 | 0.989 |
| 20 | 5 | 2000 | k-means | 0.667 | 0.444 | 0.989 |
| 20 | 6 | 2000 | FindPart-1 | 0.222 | 0.250 | 0.962 |
| 20 | 6 | 2000 | FindPart-20 | 0.222 | 0.250 | 0.962 |
| 20 | 6 | 2000 | FindPart-50 | 0.222 | 0.250 | 0.962 |
| 20 | 6 | 2000 | k-means | 0.222 | 0.250 | 0.962 |
| 20 | 7 | 2000 | FindPart-1 | 0.529 | 1.000 | 0.956 |
| 20 | 7 | 2000 | FindPart-20 | 0.529 | 1.000 | 0.956 |
| 20 | 7 | 2000 | FindPart-50 | 0.529 | 1.000 | 0.956 |
| 20 | 7 | 2000 | k-means | 0.529 | 1.000 | 0.956 |
| 20 | 8 | 2000 | FindPart-1 | 0.700 | 0.875 | 0.984 |
| 20 | 8 | 2000 | FindPart-20 | 0.700 | 0.875 | 0.984 |
| 20 | 8 | 2000 | FindPart-50 | 0.700 | 0.875 | 0.984 |
| 20 | 8 | 2000 | k-means | 0.700 | 0.875 | 0.984 |
| 20 | 9 | 2000 | FindPart-1 | 1.000 | 0.750 | 1.000 |
| 20 | 9 | 2000 | FindPart-20 | 1.000 | 0.750 | 1.000 |
| 20 | 9 | 2000 | FindPart-50 | 1.000 | 0.750 | 1.000 |
| 20 | 9 | 2000 | k-means | 0.778 | 0.875 | 0.989 |
| 20 | 10 | 2000 | FindPart-1 | 0.615 | 1.000 | 0.973 |
| 20 | 10 | 2000 | FindPart-20 | 0.615 | 1.000 | 0.973 |
| 20 | 10 | 2000 | FindPart-50 | 0.615 | 1.000 | 0.973 |
| 20 | 10 | 2000 | k-means | 0.615 | 1.000 | 0.973 |
| 25 | 1 | 2500 | FindPart-1 | 0.667 | 0.889 | 0.986 |
| 25 | 1 | 2500 | FindPart-20 | 0.667 | 0.889 | 0.986 |
| 25 | 1 | 2500 | FindPart-50 | 0.667 | 0.889 | 0.986 |
| 25 | 1 | 2500 | k-means | 0.615 | 0.889 | 0.983 |
| 25 | 2 | 2500 | FindPart-1 | 0.188 | 0.300 | 0.955 |
| 25 | 2 | 2500 | FindPart-20 | 0.278 | 0.500 | 0.955 |
| 25 | 2 | 2500 | FindPart-50 | 0.235 | 0.400 | 0.955 |
| 25 | 2 | 2500 | k-means | 0.154 | 0.200 | 0.962 |
| 25 | 3 | 2500 | FindPart-1 | 0.467 | 0.700 | 0.972 |
| 25 | 3 | 2500 | FindPart-20 | 0.467 | 0.700 | 0.972 |
| 25 | 3 | 2500 | FindPart-50 | 0.467 | 0.700 | 0.972 |
| 25 | 3 | 2500 | k-means | 0.875 | 0.700 | 0.997 |
| 25 | 4 | 2500 | FindPart-1 | 0.524 | 1.000 | 0.965 |
| 25 | 4 | 2500 | FindPart-20 | 0.524 | 1.000 | 0.965 |
| 25 | 4 | 2500 | FindPart-50 | 0.524 | 1.000 | 0.965 |
| 25 | 4 | 2500 | k-means | 0.467 | 0.636 | 0.972 |
| 25 | 5 | 2500 | FindPart-1 | 0.429 | 0.750 | 0.958 |
| 25 | 5 | 2500 | FindPart-20 | 0.429 | 0.750 | 0.958 |
| 25 | 5 | 2500 | FindPart-50 | 0.429 | 0.750 | 0.958 |
| 25 | 5 | 2500 | k-means | 0.364 | 0.333 | 0.976 |
| 25 | 6 | 2500 | FindPart-1 | 0.688 | 1.000 | 0.983 |
| 25 | 6 | 2500 | FindPart-20 | 0.688 | 1.000 | 0.983 |
| 25 | 6 | 2500 | FindPart-50 | 0.688 | 1.000 | 0.983 |
| 25 | 6 | 2500 | k-means | 0.647 | 1.000 | 0.979 |
| 25 | 7 | 2500 | FindPart-1 | 0.333 | 0.333 | 0.979 |
| 25 | 7 | 2500 | FindPart-20 | 0.400 | 0.444 | 0.979 |
| 25 | 7 | 2500 | FindPart-50 | 0.400 | 0.444 | 0.979 |
| 25 | 7 | 2500 | k-means | 0.778 | 0.778 | 0.993 |
| 25 | 8 | 2500 | FindPart-1 | 0.368 | 0.636 | 0.958 |
| 25 | 8 | 2500 | FindPart-20 | 0.263 | 0.455 | 0.952 |
| 25 | 8 | 2500 | FindPart-50 | 0.263 | 0.455 | 0.952 |
| 25 | 8 | 2500 | k-means | 0.263 | 0.455 | 0.952 |
| 25 | 9 | 2500 | FindPart-1 | 0.222 | 0.333 | 0.951 |
| 25 | 9 | 2500 | FindPart-20 | 0.208 | 0.417 | 0.934 |
| 25 | 9 | 2500 | FindPart-50 | 0.208 | 0.417 | 0.934 |
| 25 | 9 | 2500 | k-means | 0.278 | 0.417 | 0.955 |
| 25 | 10 | 2500 | FindPart-1 | 0.600 | 0.600 | 0.986 |
| 25 | 10 | 2500 | FindPart-20 | 0.636 | 0.700 | 0.986 |
| 25 | 10 | 2500 | FindPart-50 | 0.636 | 0.700 | 0.986 |
| 25 | 10 | 2500 | k-means | 0.636 | 0.700 | 0.986 |
| 30 | 1 | 3000 | FindPart-1 | 0.643 | 0.692 | 0.988 |
| 30 | 1 | 3000 | FindPart-20 | 0.714 | 0.769 | 0.991 |
| 30 | 1 | 3000 | FindPart-50 | 0.714 | 0.769 | 0.991 |
| 30 | 1 | 3000 | k-means | 0.722 | 1.000 | 0.988 |
| 30 | 2 | 3000 | FindPart-1 | 0.529 | 0.818 | 0.981 |
| 30 | 2 | 3000 | FindPart-20 | 0.529 | 0.818 | 0.981 |
| 30 | 2 | 3000 | FindPart-50 | 0.529 | 0.818 | 0.981 |
| 30 | 2 | 3000 | k-means | 0.625 | 0.909 | 0.986 |
| 30 | 3 | 3000 | FindPart-1 | 0.346 | 0.818 | 0.960 |
| 30 | 3 | 3000 | FindPart-20 | 0.357 | 0.909 | 0.958 |
| 30 | 3 | 3000 | FindPart-50 | 0.357 | 0.909 | 0.958 |
| 30 | 3 | 3000 | k-means | 0.355 | 1.000 | 0.953 |
| 30 | 4 | 3000 | FindPart-1 | 0.478 | 0.786 | 0.971 |
| 30 | 4 | 3000 | FindPart-20 | 0.478 | 0.786 | 0.971 |
| 30 | 4 | 3000 | FindPart-50 | 0.478 | 0.786 | 0.971 |
| 30 | 4 | 3000 | k-means | 0.478 | 0.786 | 0.971 |
| 30 | 5 | 3000 | FindPart-1 | 0.524 | 0.733 | 0.976 |
| 30 | 5 | 3000 | FindPart-20 | 0.524 | 0.733 | 0.976 |
| 30 | 5 | 3000 | FindPart-50 | 0.524 | 0.733 | 0.976 |
| 30 | 5 | 3000 | k-means | 0.524 | 0.733 | 0.976 |
| 30 | 6 | 3000 | FindPart-1 | 0.609 | 1.000 | 0.979 |
| 30 | 6 | 3000 | FindPart-20 | 0.609 | 1.000 | 0.979 |
| 30 | 6 | 3000 | FindPart-50 | 0.609 | 1.000 | 0.979 |
| 30 | 6 | 3000 | k-means | 0.667 | 0.571 | 0.990 |
| 30 | 7 | 3000 | FindPart-1 | 0.429 | 0.818 | 0.972 |
| 30 | 7 | 3000 | FindPart-20 | 0.429 | 0.818 | 0.972 |
| 30 | 7 | 3000 | FindPart-50 | 0.429 | 0.818 | 0.972 |
| 30 | 7 | 3000 | k-means | 0.533 | 0.727 | 0.983 |
| 30 | 8 | 3000 | FindPart-1 | 0.688 | 0.786 | 0.988 |
| 30 | 8 | 3000 | FindPart-20 | 0.688 | 0.786 | 0.988 |
| 30 | 8 | 3000 | FindPart-50 | 0.688 | 0.786 | 0.988 |
| 30 | 8 | 3000 | k-means | 0.688 | 0.786 | 0.988 |
| 30 | 9 | 3000 | FindPart-1 | 0.273 | 0.545 | 0.962 |
| 30 | 9 | 3000 | FindPart-20 | 0.316 | 0.545 | 0.969 |
| 30 | 9 | 3000 | FindPart-50 | 0.316 | 0.545 | 0.969 |
| 30 | 9 | 3000 | k-means | 0.429 | 0.545 | 0.981 |
| 30 | 10 | 3000 | FindPart-1 | 0.304 | 0.700 | 0.962 |
| 30 | 10 | 3000 | FindPart-20 | 0.412 | 0.700 | 0.976 |
| 30 | 10 | 3000 | FindPart-50 | 0.412 | 0.700 | 0.976 |
| 30 | 10 | 3000 | k-means | 0.412 | 0.700 | 0.976 |
| 35 | 1 | 3500 | FindPart-1 | 0.632 | 0.857 | 0.988 |
| 35 | 1 | 3500 | FindPart-20 | 0.632 | 0.857 | 0.988 |
| 35 | 1 | 3500 | FindPart-50 | 0.632 | 0.857 | 0.988 |
| 35 | 1 | 3500 | k-means | 0.706 | 0.857 | 0.991 |
| 35 | 2 | 3500 | FindPart-1 | 0.423 | 0.917 | 0.974 |
| 35 | 2 | 3500 | FindPart-20 | 0.423 | 0.917 | 0.974 |
| 35 | 2 | 3500 | FindPart-50 | 0.423 | 0.917 | 0.974 |
| 35 | 2 | 3500 | k-means | 0.500 | 0.917 | 0.981 |
| 35 | 3 | 3500 | FindPart-1 | 0.476 | 0.714 | 0.981 |
| 35 | 3 | 3500 | FindPart-20 | 0.474 | 0.643 | 0.983 |
| 35 | 3 | 3500 | FindPart-50 | 0.474 | 0.643 | 0.983 |
| 35 | 3 | 3500 | k-means | 0.500 | 0.714 | 0.983 |
| 35 | 4 | 3500 | FindPart-1 | 0.476 | 0.625 | 0.981 |
| 35 | 4 | 3500 | FindPart-20 | 0.458 | 0.688 | 0.978 |
| 35 | 4 | 3500 | FindPart-50 | 0.458 | 0.688 | 0.978 |
| 35 | 4 | 3500 | k-means | 0.421 | 0.500 | 0.981 |
| 35 | 5 | 3500 | FindPart-1 | 0.462 | 0.750 | 0.976 |
| 35 | 5 | 3500 | FindPart-20 | 0.455 | 0.938 | 0.969 |
| 35 | 5 | 3500 | FindPart-50 | 0.455 | 0.938 | 0.969 |
| 35 | 5 | 3500 | k-means | 0.455 | 0.938 | 0.969 |
| 35 | 6 | 3500 | FindPart-1 | 0.280 | 0.538 | 0.969 |
| 35 | 6 | 3500 | FindPart-20 | 0.308 | 0.615 | 0.969 |
| 35 | 6 | 3500 | FindPart-50 | 0.348 | 0.615 | 0.974 |
| 35 | 6 | 3500 | k-means | 0.310 | 0.692 | 0.966 |
| 35 | 7 | 3500 | FindPart-1 | 0.636 | 0.824 | 0.986 |
| 35 | 7 | 3500 | FindPart-20 | 0.632 | 0.706 | 0.988 |
| 35 | 7 | 3500 | FindPart-50 | 0.632 | 0.706 | 0.988 |
| 35 | 7 | 3500 | k-means | 0.588 | 0.588 | 0.988 |
| 35 | 8 | 3500 | FindPart-1 | 0.571 | 0.750 | 0.984 |
| 35 | 8 | 3500 | FindPart-20 | 0.500 | 0.563 | 0.984 |
| 35 | 8 | 3500 | FindPart-50 | 0.500 | 0.563 | 0.984 |
| 35 | 8 | 3500 | k-means | 0.571 | 0.750 | 0.984 |
| 35 | 9 | 3500 | FindPart-1 | 0.356 | 1.000 | 0.950 |
| 35 | 9 | 3500 | FindPart-20 | 0.341 | 0.938 | 0.950 |
| 35 | 9 | 3500 | FindPart-50 | 0.469 | 0.938 | 0.971 |
| 35 | 9 | 3500 | k-means | 0.429 | 0.750 | 0.972 |
| 35 | 10 | 3500 | FindPart-1 | 0.455 | 0.625 | 0.979 |
| 35 | 10 | 3500 | FindPart-20 | 0.524 | 0.688 | 0.983 |
| 35 | 10 | 3500 | FindPart-50 | 0.524 | 0.688 | 0.983 |
| 35 | 10 | 3500 | k-means | 0.545 | 0.750 | 0.983 |

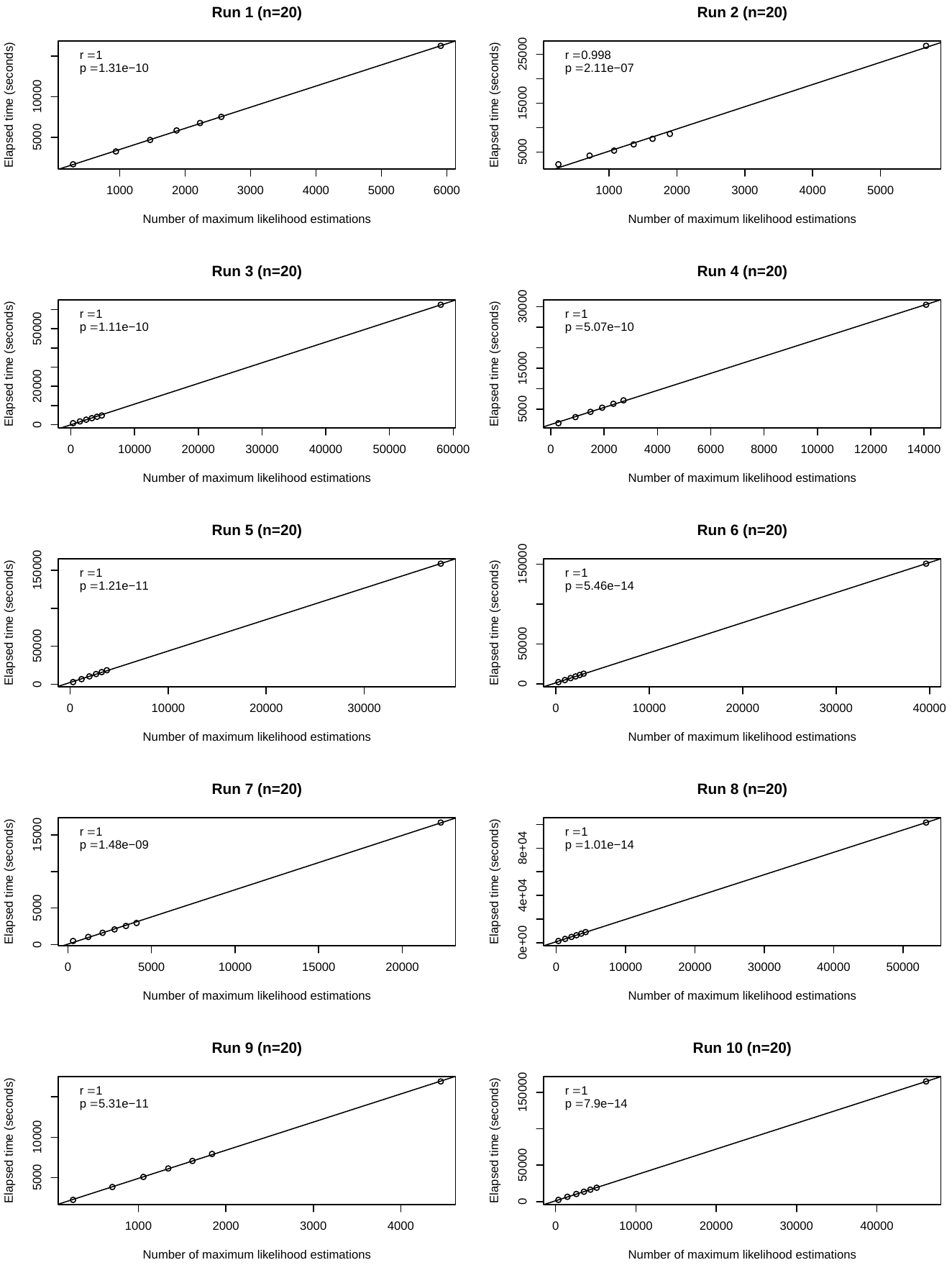
**Fig S1. Correlation between the number of the maximum likelihood estimations and the total elapsed time of FindPart-**$\boldsymbol{w}$ **for** $\boldsymbol{n}\mathbf{=20}$**. The elapsed time did not include the time to calculate of 95% confidence intervals of parameters. The** $\boldsymbol{x}$**-axis shows the number of the maximum likelihood estimations and the** $\boldsymbol{y}$**-axis shows the total running time of FindPart-**$\boldsymbol{w}$ **for** $\boldsymbol{n}\mathbf{=20}$**. r is the correlation coefficient, and p is its corresponding p-value.**
